# Systematic analysis of snRNA genes reveals frequent *RNU2-2* variants in dominant and recessive developmental and epileptic encephalopathies

**DOI:** 10.1101/2025.09.02.25334923

**Authors:** Elsa Leitão, Amandine Santini, Benjamin Cogne, Myriam Essid, Maria Athanasiadou, Christy W. LaFlamme, Pierre Marijon, Virginie Bernard, Nicolas Chatron, Giulia Barcia, Boris Keren, Cyril Mignot, Perrine Charles, Thomas Besnard, Jean-Madeleine de Sainte Agathe, Edith P. Almanza Fuerte, Soham Sengupta, Mathieu Milh, Francis Ramond, Talia Allan, Isabelle An, Camila Araujo, Stephanie Arpin, Christina Austin-Tse, Stéphane Auvin, Sarah Baer, Nadia Bahi-Buisson, Mads Bak, Magalie Barth, Stéphanie Baulac, Nathalie Bednark Weirauch, Matthias Begemann, Mark F. Bennett, Uriel Bensabath, Stéphane Bézieau, Rakia Bhouri, Margaux Biehler, Trine Bjørg Hammer, Julie Bogoin, Emilie Bonanno, Simon Boussion, Nuria C. Bramswig, Céline Bris, Adelaide Brosseau-Beauvir, Ange-Line Bruel, Julien Buratti, Pascal Chambon, Nicole Chemaly, Bertrand Chesneau, Estelle Colin, Maxime Colmard, Solène Conrad, Thomas Courtin, Louis T. Dang, Anne de Saint Martin, Caroline de Vanssay de Blavous Legendre, Anne-Sophie Denommé-Pichon, Stephanie DiTroia, Martine Doco-Fenzy, Christèle Dubourg, Charlotte Dubucs, Stéphanie Ducreux, Louis Dufour, Romain Duquet, Benjamin Durand, Salima El Chehadeh, Miriam Elbracht, Laurence Faivre, Marie Faoucher, Anne Faudet, Sylvie Forlani, Mélanie Fradin, Pauline Gaignard, Benjamin Ganne, Aurore Garde, Justine Géraud, Deepak Gill, Alice Goldenberg, David Grabli, Coraline Grisel, Sophie Gueden, Paul Gueguen, Anne-Marie Guerrot, Agnès Guichet, Nina Härting, Martin Georg Häusler, Solveig Heide, Bénédicte Héron, Delphine Héron, Mathilde Heulin, Clara Houdayer, Bertrand Isidor, Aurélia Jacquette, Louis Januel, Nolwenn Jean-Marçais, Kevin Jousselin, Frank J. Kaiser, Sabine Kaya, Chontelle King, Marina Konyukh, Florian Kraft, Jeremias Krause, Rémi Kirstetter, Alma Kuechler, Ingo Kurth, Audrey Labalme, Jean-Serene Laloy, Vincent Laugel, Floriane Le Bricquir, Anne-Sophie Lèbre, Marine Lebrun, Eric Leguern, Jonathan Levy, Nico Lieffering, Stanislas Lyonnet, Kevin Lüthy, Sian Macdonald, Lamisse Mansour-Hendili, Julien Maraval, Carolin Mattausch, Olfa Messaoud, Godelieve Morel, Jérémie Mortreux, Arnold Munnich, Rima Nabbout, Sophie Nambot, Vincent Navarro, Ashana Neale, Laetitia Nguyen, Mathilde Nizon, Frédérique Nowak, Melanie C. O’Leary, Sylvie Odent, Naomi Meave Ojeda, Valerie Olin, Katrin Õunap, Lynn S. Pais, Robin Paluch, Eleni Panagiotakaki, Olivier Patat, Laurence Perrin-Sabourin, Florence Petit, Christophe Philippe, Amélie Piton, Marc Planes, Céline Poirsier, Antoine Pouzet, Clément Prouteau, Sylvia Quéméner-Redon, Mathilde Renaud, Anne-Claire Richard, Marlène Rio, Clotilde Rivier, Florence Robin-Renaldo, Paul Rollier, Massimiliano Rossi, Agathe Roubertie, Mailys Rupin, Pascale Saugier-Veber, Russell Saneto, Elisabeth Sarrazin, Elise Schaefer, Caroline Schluth-Bolard, Amy Schneider, Isabell Schumann, Vladimir Seplyarskiy, Thomas Smol, Shamil Sunyaev, Brian Sperelakis-Beedham, Sarah L. Stenton, Friedrich Stock, Mylene Tharreau, Deniz Torun, Joseph Toulouse, Harshini Thiyagarajah, Stéphanie Valence, Sophie Valleix, Laurent Villard, Dorothée Ville, Nathalie Villeneuve, Antonio Vitobello, Aurélie Waernessyckle, Yvonne Weber, Dagmar Wieczorek, Tom Witkowski, Manya Yadavilli, Tony Yammine, Khaoula Zaafrane-Khachnaoui, Maha S. Zaki, Alban Ziegler, Alban Lermine, Gael Nicolas, Joseph G. Gleeson, Lynette G. Sadleir, Michael S. Hildebrand, Ingrid E. Scheffer, Nicola Whiffin, Anne O’Donnell-Luria, Heather C. Mefford, Pierre Blanc, Julien Thevenon, Camille Charbonnier, Clément Charenton, Christel Depienne, Gaetan Lesca, Caroline Nava

## Abstract

Variants in spliceosomal small nuclear RNA (snRNA) genes *RNU4-2* (ReNU syndrome), *RNU5B-1*, and *RNU2-2* have recently been linked to dominant neurodevelopmental disorders (NDDs), revealing a major, previously overlooked role for noncoding snRNAs in human disease. Here, we systematically analysed 200 potentially functional snRNA genes in a French cohort comprising 26,911 individuals with rare disorders and through international collaborations. We identify *de novo* and biallelic variants in *RNU2-2* associated with both dominant and recessive NDDs in 126 individuals from 108 unrelated families. Recessive *RNU2-2* NDD is at least twice as frequent as the dominant NDD caused by n.4G>A and n.35A>G, and often arises from a *de novo* variant in *trans* with an inherited allele, reflecting the high mutability of snRNA genes. Dominant and recessive *RNU2-2*-NDDs share overlapping clinical features with frequent epilepsy. Blood transcriptomics and DNA methylation analyses revealed subtle, variant-specific effects on splicing and episignatures. Our findings support a gradient-of-impact model and a continuum between dominant and recessive inheritance, establishing *RNU2-2* variants as a frequent cause of NDDs, nearly as prevalent as ReNU syndrome.

## Introduction

Small nuclear RNAs (snRNAs) are non-coding RNAs essential for RNA processing, particularly splicing of pre-messenger RNAs (mRNA). The spliceosome, which is a dynamic ribonucleoprotein (RNP) that catalyses splicing, relies on five uridine-rich snRNAs for its assembly and function. In mammals, two spliceosome types operate depending on the intron class: the major spliceosome processes >99% of introns with GU-AG splice sites (U2-type) using snRNAs U1, U2, U4, U5, and U6, and the minor spliceosome excises rare AU-AC introns (U12-type) with snRNAs U11, U12, U4atac, and U6atac, with U5 being shared by both complexes.^1,2^

Minor spliceosome snRNA genes are typically single-copy, whereas major spliceosome genes have multiple functional copies. Human genomes contain at least two U4, five U5, and five U6 functional genes. U1 genes include at least four identical copies (*RNU1-1* to *RNU1-4*) on chromosome 1p36.13 and over 30 ‘variant’ U1 genes (*RNVU1*) on chromosome 1q21.1-q23.3^3-5^. Most genes encoding U2 are organized in large tandem arrays on chromosome 17q21.31, with copy numbers ranging from 6 to >80 copies per chromosome^6^.

Biallelic pathogenic variants were first described in *RNU4ATAC* and cause phenotypically variable developmental disorders: microcephalic osteodysplastic primordial dwarfism type I, Taybi-Linder, Lowry-Wood, or Roifman syndromes^7-11^. Recessive variants in *RNU12* may lead to craniosynostosis, anal anomalies and skin lesions (CDAG) and/or spinocerebellar ataxia^12,13^ although definitive evidence is missing.

In 2024, recurrent d*e novo* variants in the major spliceosome gene *RNU4-2* were initially shown to cause ReNU syndrome (OMIM 620851), one of the most common known neurodevelopmental disorders (NDDs).^14,15^ These dominant variants are located within an 18-bp critical region spanning the T loop and part of the stem III, facing the U6 ACAGAGA box and enabling 5′ splice site recognition^14,16^. *RNU4-2* pathogenic variants are associated with mild but specific, widespread splicing and methylation abnormalities in blood cells of affected individuals, the degree of which correlates with disease severity^14,16^. *De novo* variants in *RNU5B-1* (and possibly *RNU5A-1*) clustering in the U5 5’ loop I lead to NDD with variable malformations^16,17^. The recent discovery of biallelic loss-of-function variants in other regions of *RNU4-2* expanded its mutational spectrum and added complexity to variant interpretation.^18,19^

Identifying variants in snRNA genes is further complicated by sequence redundancy and incomplete annotations, especially when distinguishing functional genes from pseudogenes. Notably, *de novo* pathogenic variants in *RNU2-2P*, initially annotated as a pseudogene, were recently linked to a dominant NDD with epilepsy^17,20^. Expression analysis revealed levels comparable to canonical U2 genes^21^, leading to its reclassification as *RNU2-2*. Unlike *RNU4-2*, no detectable splicing anomalies were found in blood transcriptomes of patients with *RNU2-2* variants^20^. Strikingly, active snRNA and tRNA genes accumulate non-pathogenic variants up to 10 times faster than other regions, a feature that may help distinguish functional snRNA genes from pseudogenes.^22^

In this study, we build on our previous work involving 50 Genome Organisation (HUGO) Gene Nomenclature Committee (HGNC)-approved snRNA genes,^16^ extending it to systematically investigate variants in possibly functional snRNA genes in a large French cohort with rare diseases. This led us to report a highly prevalent recessive NDD with epilepsy caused by biallelic variants in *RNU2-2* that was validated through international collaborations.

## Results

### Identification of potentially functional spliceosomal snRNA genes

To distinguish functional snRNA genes from pseudogenes, we performed an *in silico* analysis of all annotated snRNA genes (Methods). The Ensembl database contains 2,094 genes encoding snRNAs in the hg38 reference genome. Of these, 1,741 snRNAs are components of the spliceosome, while the remainder have other functions or are not located on identifiable chromosomes. We prioritized genes located in open chromatin regions marked by H3K27 acetylation histone marks (H3K27ac) in the embryonic stem cell line H1-hESC, containing proximal cis-regulatory elements (cCREs; from ENCODE^23^), or previously reported as hypermutable.^22^ HGNC-approved genes were also included^16^. This analysis yielded 200 potentially functional snRNA genes, including 147 ‘pseudogenes’ and *RNU2-2* (Fig. 1a; Supplementary Table 1). The breakdown of snRNA types aligns with prior observations: minor spliceosome snRNAs exist in few copies (range 1-3), whereas major spliceosome snRNAs are present in many more copies (range 11-117), and U5, shared by both complexes, is intermediate (*n*=9; Fig. 1b-c). U6 has the highest number of copies.

**Fig 1.**
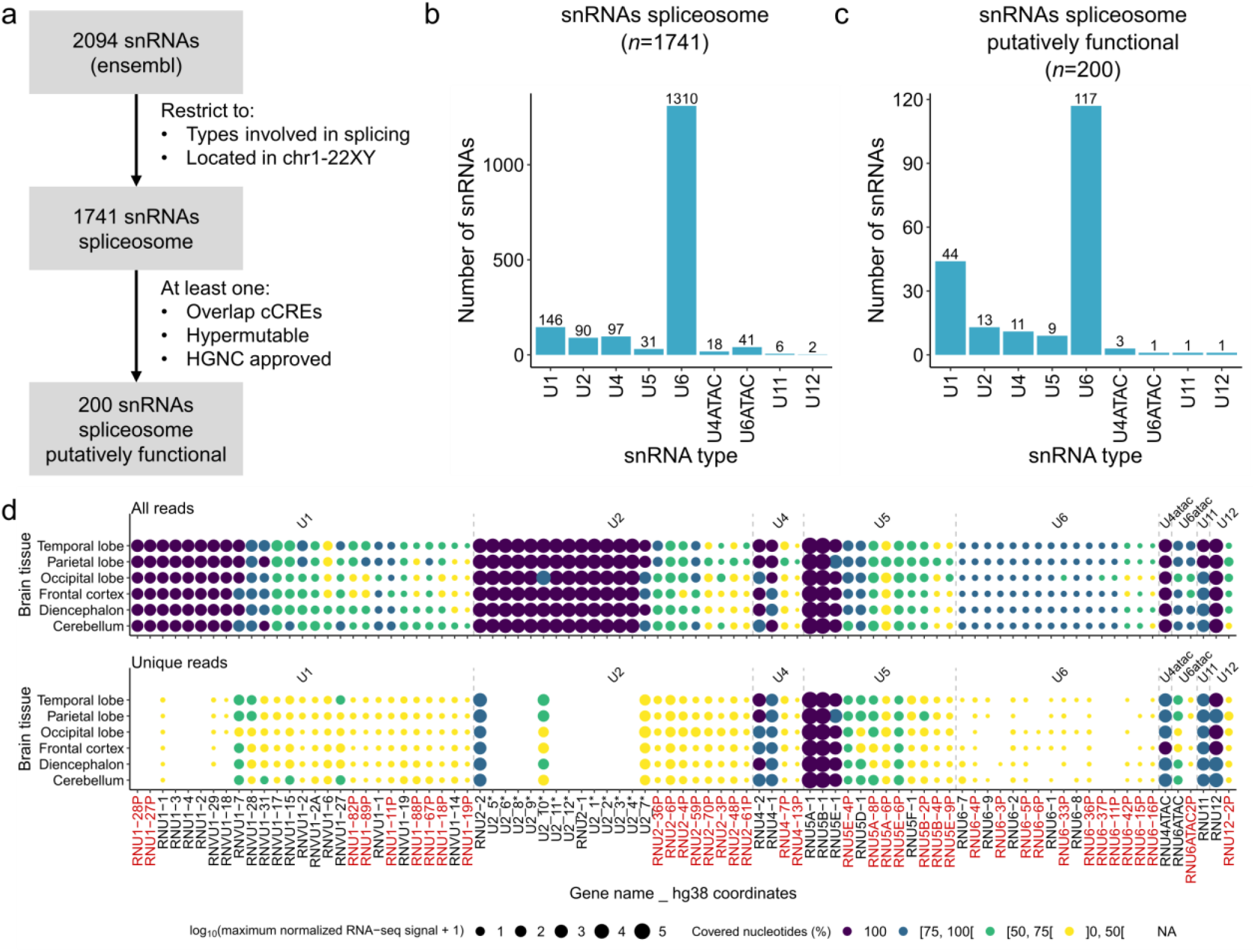
Systematic *in silico* analysis of possible functional snRNAs. **a**, Filtering strategy used to retain genes encoding possible functional snRNAs. **b**, and **c**, Number and distribution of annotated spliceosomal snRNAs before (**b**) and after (**c**) filtering **d**, Expression of snRNAs in the human brain from ENCODE small RNA-seq data. Upper panel: all mapped reads (including multi-mapped). Lower panel: uniquely mapped reads. Expression is shown as log10 of the maximum normalized RNA-seq signal. Colours indicate the proportion of the snRNA length covered by mapped reads: 100% (purple), 75-99% (dark blue), 50-74% (green), <50% (yellow).

In parallel, we reanalysed an ENCODE brain small RNA-seq dataset.^24^ We performed two analyses: one using all mapped reads, including multi-mapped, and another restricted to uniquely mapped reads. The first captures expression from all identical copies, while the second reflects only uniquely assignable snRNAs. In total, 87 putatively functional snRNAs, including 39 annotated as pseudogenes, were detectable in the human brain (Fig. 1d).

### Analysis of *de novo* and biallelic variants in putatively functional snRNA genes in the PFMG cohort

We next analysed variants in the 200 potentially functional snRNA genes in the Plan France Médecine Génomique 2025 (PFMG) cohort,^25^ which at the time comprised short-read genome data from 26,911 patients with rare disorders (18,186 with NDDs). We focused on *de novo* and biallelic variants, hypothesising roles in dominant or recessive monogenic disorders.

A total of 6,971 variants (4,168 *de novo* and 4,427 biallelic) in 101 genes were identified in SeqOIA and 1,946 variants (511 *de novo* and 1,583 biallelic) in 75 genes were identified in Auragen, across a total of 5,222 patients (Ext Data Fig. 1; Supplementary Fig. 1). Analysis of variant allele fractions (VAF) for variants covered by ≥10 reads showed that many snRNA gene variants fall in the low range of 0.1–0.3. This pattern, seen in both sub-cohorts, reflects the limitation of short-read sequencing in distinguishing identical or highly similar snRNA copies. Recurrent low VAFs were observed in *RNU1-1, RNU1-2, RNU2-1*, other closely related U1 and U2 copies, and several U6 genes, consistent with their high sequence identity (Ext. Data Fig. 2; Supplementary Fig. 2).To address this, we excluded 43 genes: 10 with predominantly low-VAF variants and 39 (30 with variants) overlapping >50% with the GIAB Problematic Regions UCSC track. Following this filtering, we retained a total of 893 high-quality variants in 81 genes, including 194 *de novo* and 761 biallelic variants, across 808 patients in the entire PFMG cohort.

**Fig 2.**
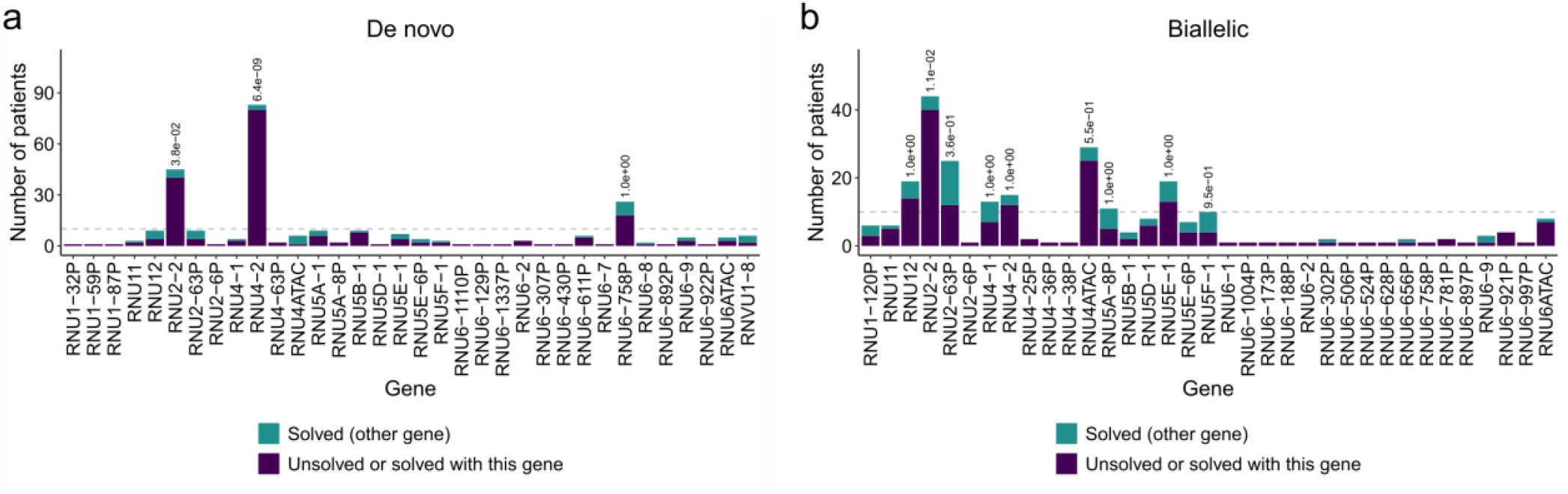
Identification of potential novel snRNA gene–disease associations in the PFMG cohort. The cohort was divided into solved (*n*=8,343) and unsolved (*n*=18,568) cases for discovery analyses, with cases solved by variants in snRNAs with known disease association merged into the unsolved group. We compared the proportion of cases with rare variants (gnomAD allele count < 100) between solved and unsolved groups for rare *de novo* variants (**a**) and rare biallelic variants (**b**). Fisher’s test was used to test statistical enrichment in unsolved versus solved cases for genes in which at least 10 patients had variants (minimum number needed to reach statistical significance in the cohort). *P*-values are shown above the bars.

To identify possible new disease gene associations, we divided the cohort into solved (*n*=8,343) and unsolved (*n*=18,568) cases for subsequent analyses. Since pathogenic variants in four snRNA genes already associated with disease (*RNU4ATAC, RNU4-2, RNU5B-1, RNU2-2*) were present in the dataset, we manually curated these cases and aggregated them with the unsolved category, as they would have been prior to the gene-disease association. Focusing on rare variants (allele counts <100 in gnomAD_v3, as defined by the distribution observed in our cohort; Ext Data Fig. 3; Supplementary Fig. 3), we compared *de novo* variants in 38 genes between solved and unsolved cases (or cases solved by the mentioned snRNA genes), finding a significant enrichment in only *RNU4-2* and *RNU2-2* (Fig. 2a; Supplementary Fig. 4a-b; Supplementary Fig. 5a-c). For biallelic variants, using the same AC <100 filter, we identified a significant enrichment in *RNU2-2* in the combined cohorts (*P*=0.011; two-sided Fisher’s test) while *RNU4ATAC* reached significance only in the SeqOIA subcohort (*P*=0.012; two-sided Fisher’s test; Fig. 2b; Supplementary Fig. 4ac-d; Supplementary Fig. 5d-f).

**Fig 3.**
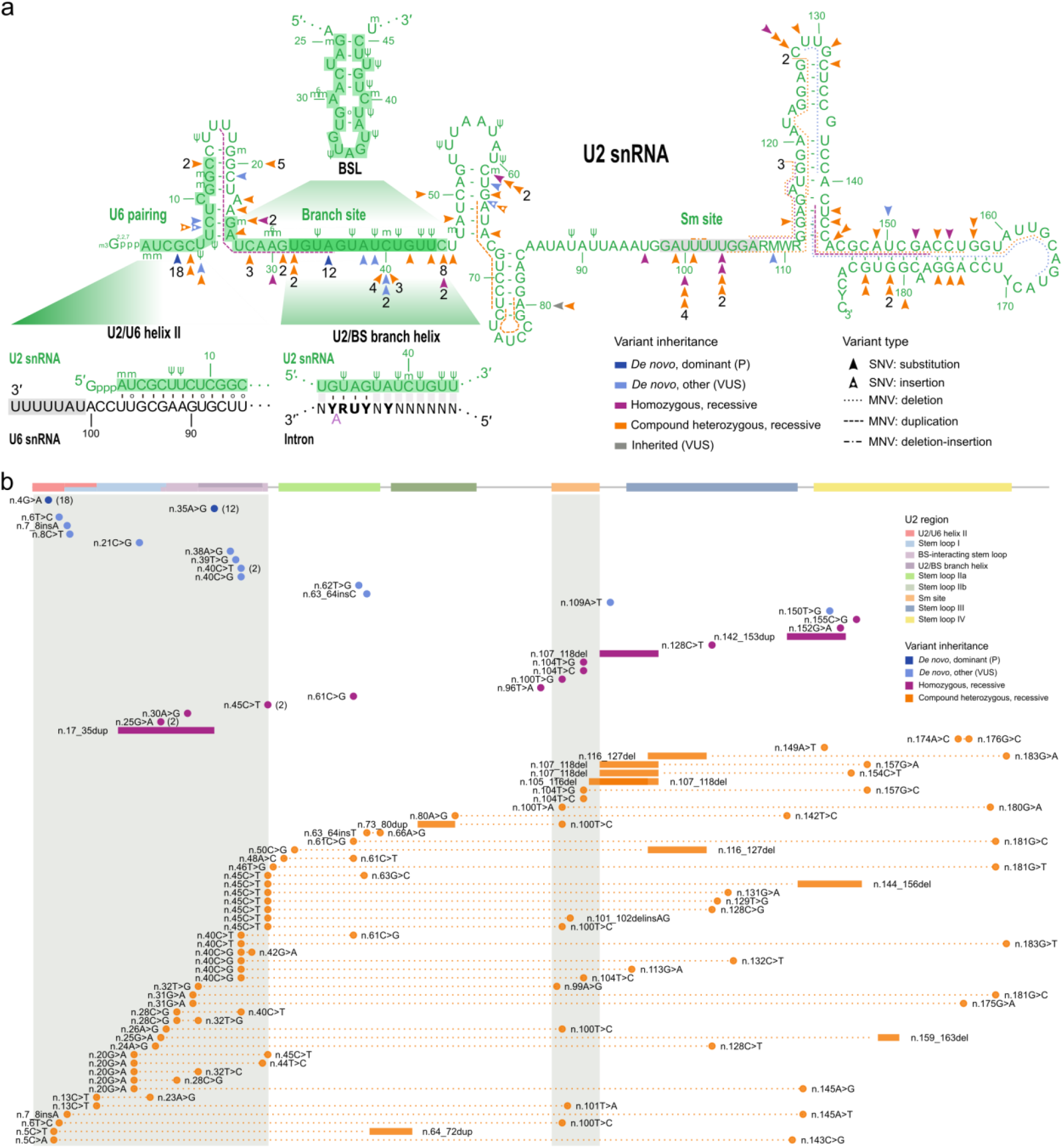
Overview of *RNU2-2* variants identified in this study. **a**, Two-dimensional predicted structure of U2-2 snRNA showing structural and functional domains. Arrow heads indicate point variants identified in this study. Variants are coloured according to their inheritance: dark blue: *de novo*, dominant (n.4G>A or n.34A>G); light blue: *de novo* other (VUS); orange: compound, recessive; purple: homozygous, recessive. The numbers in black represent the count of patients with each variant, for variants identified in more than one family. Other variant types are shown with dotted (deletions), dashed (duplications) or dotted-dashed (indels) lines. The nucleotide differences between *RNU2-2* and *RNU2-1* are shown using IUPAC codes. Green numbers refer to the numbering of U2-2 nucleotides. Ψ: pseudouridine, m: 2’-O-methyl residues; m6: N6-methyladenosine; ^2,2,7^m3Gppp: 2,2,7-trimethylguanosine cap. Green-shaded regions: functional domains of U2 involved in spliceosomal activity: U2/U6 helix II (nt 1-13); branch-point-interacting stem loop (BSL; nt 25–45); and U2/BS branch helix (nt 32-44). Grey-shaded region: Sm site. **b**, Locations of *RNU2-2* variants on linear (unfolded) U2-2 snRNA, showing clustering of variants in the U2/U6 helix, BSL regions, and Sm site (grey-shaded), as well as preferential associations of compound heterozygous variants. Variants are ordered by inheritance mode and by the position of the first variant on the snRNA. Variant colouring is the same as in Fig 3a.

### *RNU2-2* variants as a cause of dominant and recessive disorders

Our findings suggest that *RNU2-2* variants underlie both dominant and recessive disorders, consistent with recent observations for *RNU4-2*.^18,19^ We reanalysed the PFMG cohort using relaxed criteria filtering solely on homozygous variants. Given the high *de novo* rate and variant burden in *RNU2-2*, we transitioned to the *All of Us* database to filter potentially pathogenic variants (Methods; Supplementary Fig. 6). Criteria were allele count (AC) < 50 for *de novo* variants and AC < 200 for biallelic variants. We also included three families carrying recurrent variants in *trans* with more common alleles. Additional cases were identified through international collaborations using targeted or genome sequencing.

In the PFMG cohort, 38 unrelated patients had rare *de novo RNU2-2* variants (Supplementary Table 2). Of these, 20 unrelated patients and one monozygotic twin harboured previously reported pathogenic alleles: n.4G>A (*n*=10) and n.35A>G (*n*=11 including the twin). One patient with n.35A>G was mosaic, and deep targeted sequencing (>2000×) confirmed mutant allele fractions of 12% (678/5218 reads) in blood, 20% (1181/5659 reads) in urine, and 25% (574/2267 reads) in buccal cells. The n.4G>A variant was also observed in a singleton in the PFMG cohort. Furthermore, an additional seven n.4G>A (six *de novo*, one non-maternal) and two n.35A>G variants (both *de novo*) were identified in patients from other cohorts. In total, n.4G>A and n.35A>G were identified in 18 and 13 patients, respectively, including the mosaic case and monozygotic twin

The remaining 18 patients from the PFMG cohort and two from other cohorts had *de novo* variants other than n.4G>A and n.35A>G. These included n.5C>A, n.6T>C (*n*=2), n.7_8insA (*n*=2), n.8C>T, n.21C>G, n.31G>A, n.32T>G, n.38A>G, n.40C>G, n.40C>T (*n*=3), n.62T>G, n.63_64insC, n.109A>T, n.129_139del, n.143_167del, and n.150T>G (Fig. 3). Six of these *de novo* variants (n.5C>A, n.6T>C, n.7_8insA, n.31G>A, n.32T>G, and n.40C>T) were found in individuals who also carried a second rare variant in *trans*, suggesting recessive inheritance. In contrast, among cases with n.4G>A, only one individual had a second variant in *trans* (n.80A>G; Supplementary Table 2).

In addition, we identified rare biallelic variants in 58 probands, 43 from the PFMG cohort and 15 from other cohorts. Of these, 40 carried compound heterozygous variants, 16 had homozygous variants and two harboured hemizygous variants associated *in trans* with a complete or partial gene deletion (Fig. 3; Ext Fig. 4). In 15 families, one or more siblings were also affected, with both variants co-segregating with the disease in affected siblings (Fig. 4a; Ext Data Fig. 5). Seventeen variants were recurrently found in unrelated families (n.45C>T, *n*=10; n.20G>A, *n*=5; n.40C>G, n.100T>C, n.107_118del, *n*=4 each; n.25G>A, n.28C>G, n.40C>T, n.61C>G, n.104T>C, *n*=3 each; n.13C>T, n.31G>A, n.32T>G, n.104T>G, n.116_127del, n.128C>T, n.181G>C, *n*=2 each).

**Fig 4.**
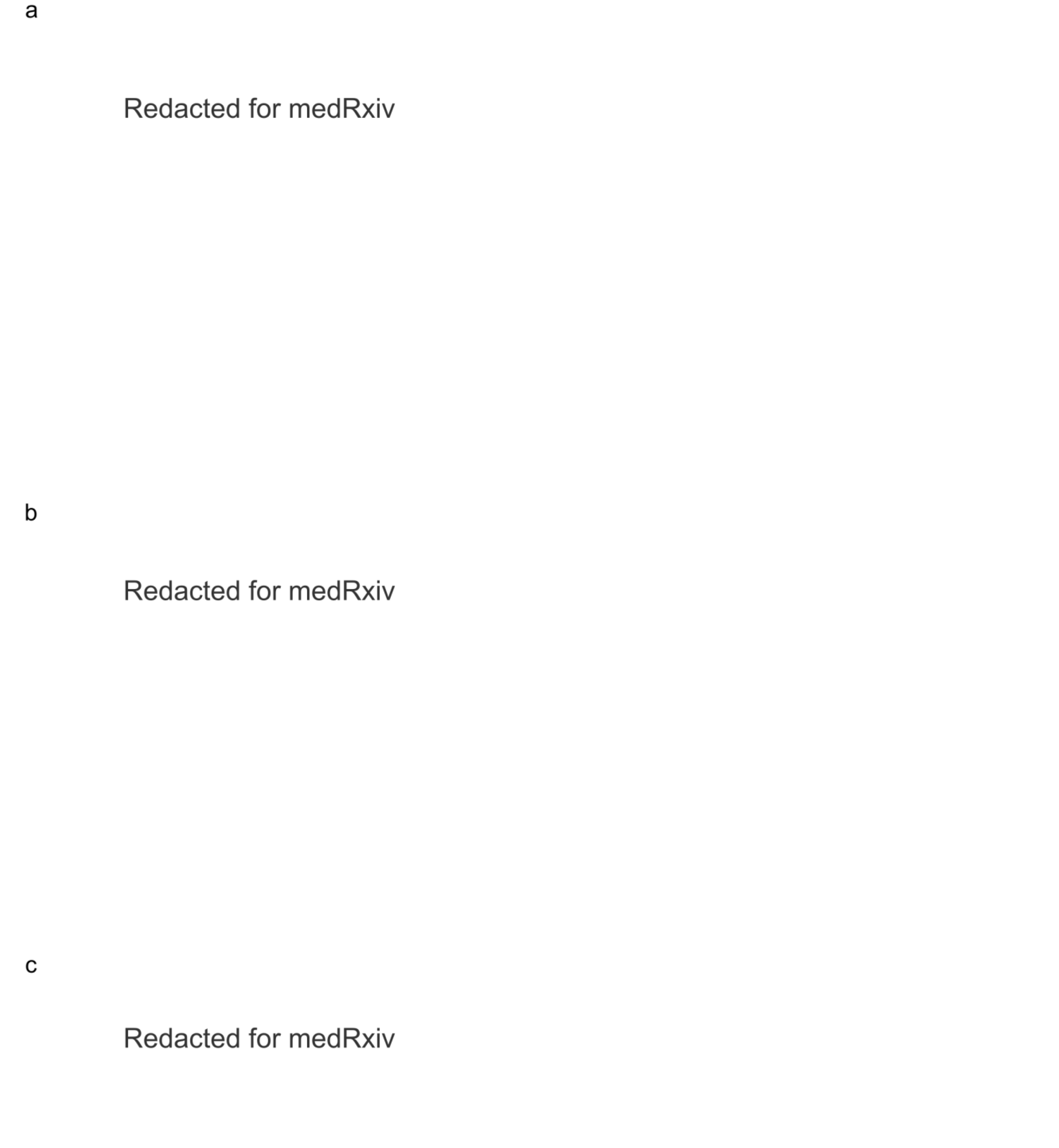
Variant segregation and facial features associated with *RNU2-2* variants. (Retracted for Med Rxiv submission)

**Fig 5.**
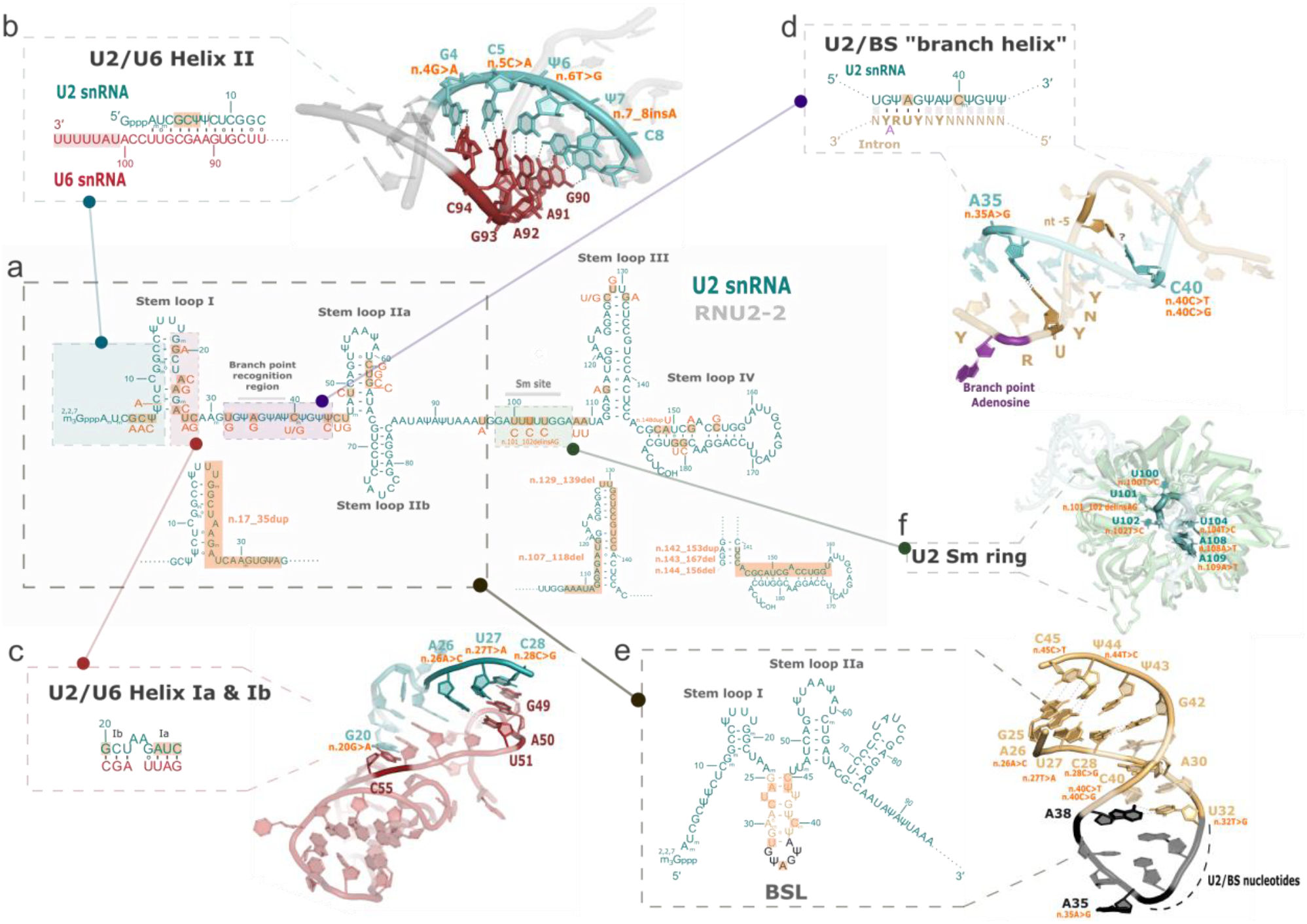
Possible functional impact of *RNU2-2* variants. Overview of *RNU2-2* variants identified in this study. **a**, Two-dimensional structure of the U2 small nuclear RNA (green). Orange boxes indicate variants from this study, with point mutations and single nucleotide insertions represented on the graph along with their respective changes. ^2,2,7^m3Gppp: 2,2,7-trimethylguanosine cap, Ψ: pseudouridine, m: 2’-O-methyl residues. **b**, Zoom-in box representing the two-dimensional predicted structure of the U2 (green) and U6 (red) small nuclear RNAs (snRNAs) U2/U6 helix II during formation of precatalytic spliceosome. Interactions stabilizing these structures as well as mutations potentially affecting their stability are represented (PDB 5XJC). Shaded region: Lsm/Sm protein binding site of U6 snRNA. **c**, Zoom-in box representing the two-dimensional predicted structure of U2/U6 helixes Ia and Ib. Nucleotides involved in the structure and associated with pathological variants are represented (PDB 5XJC). **d**, Close-up of U2/BS branch helix where the two-dimensional predicted structure of the interaction between branch point recognition region of U2 snRNA and the intronic branch site (light brown) is represented. The branch point adenosine is highlighted in purple. ‘YNYURAY’: consensus of branch site in metazoans. The mutations related with this region are depicted on the structure (PDB 5XJC). **e**, Zoom-in box representing the two-dimensional predicted structure of 17S U2 snRNA and the branch point interacting stem loop (BSL) is highlighted. In blue the adenine at position 30 that interacts with the cytosine (purple) at position 40 linked to two reported variants. In black the nucleotides involved directly on the U2/BS formation. The structure of 17S U2 snRNAs’ BSL structure is represented (PDB 7Q3L). **f**, Close-up of the U2 snRNA Sm ring (light green) where the nucleotides with the mutations close to or within it are marked in orange (PDB 5XJC).

In total, we identified 126 individuals from 108 unrelated families with *RNU2-2* variants: 31 with pathogenic dominant variants (n.4G>A and n.35A>G, including the twin); 81 patients from 64 unrelated families with biallelic variants; and 14 patients with single *de novo* heterozygous variants other than n.4G>A and n.35A>G. We identified 83 distinct variants that we classified using ACMG criteria (Supplementary Table 3). Among 102 patients with genome data, only one carried a *de novo* nonsense variant in *GATA3* partially explaining his phenotype; all others remained unsolved. Overall, 103 of 108 probands presented with NDD, while the remaining five included two terminated fetuses, one toddler with hearing impairment, and two non-NDD patients (one with the *GATA3* variant). In the PFMG cohort, NDD was the predominant presentation, observed in 79/82 patients (*P*=1.9 × 10^−20^, one-tailed binomial test; 95%; CI: 0.91–1).

Eight *de novo* variants could be reliably phased: five, including all phaseable occurrences of the pathogenic n.4G>A (*n*=3) and n.35A>G (*n*=1) (along with n.6T>C) were phased to maternal alleles, whereas three (n.40C>T, n.129_139del, and n.143_167del) occurred *de novo* on paternal alleles (Supplementary Table 2).

### Dominant and recessive *RNU2-2* NDDs share overlapping features

We next aimed to delineate the clinical spectrum associated with both dominant and recessive *RNU2-2* disorders. Detailed clinical data were collected for 79 patients (40 females, 39 males), including 17 with n.4G>A, 10 with n.35A>G, and 5 with another monoallelic *de novo* variant, as well as 47 patients with biallelic inheritance (Table 1; Supplementary Table 4). The median age at inclusion in the study was 14 years (range: 0 (fetus) - 46 years).

Overall, taking only patients with dominant pathogenic (*n*=27) and biallelic variants (*n*=47) into account, developmental delay and intellectual disability were present in all cases except two: a terminated fetus with polymalformation syndrome (n.174A>C/n.176G>C), and a patient with developmental delay without ID, but fragile visuoconstructive reasoning (n.20G>A/n.145A>G). Severe/profound intellectual disability was most frequent (54/64, 84.5%), followed by moderate (9/65, 14%) and mild (1/64, 1.5%) ID. Epilepsy occurred in 89% patients (65/73), with identical rates in dominant (24/27, 89%) and recessive (41/46, 89%) cases, and onset ranging from 8 weeks to 16 years. Among those with epilepsy, 78.5% (51/65) had seizure onset between 8 weeks and 3 years (monoallelic: 18/24; biallelic: 33/41), while 21.5% (14/65) had onset after 3 years (monoallelic: 6/24; biallelic: 8/41). Seizure types were variable. In biallelic families, 22/26 (85%) had generalized seizures, whereas in monoallelic cases, 16/23 (70%) had focal seizures, including hemicorporal seizures in 7/22 (32%). Generalized tonic-clonic seizures occurred in 28/48 patients (58%; biallelic 19/26, monoallelic 9/22), myoclonic seizures in 69% versus 27%, and epileptic spasms in 39% versus 23%. Absence seizures were reported in 26/52 patients (50%; monoallelic 10/23, biallelic 16/29), and drug resistance in 44/60 patients (73%; monoallelic 17/23, biallelic 27/37).

Clinical presentations were variable within each group (n.4G>A, n.35A>G, and biallelic variants), but overall, the clinical spectrum were broadly similar, with no striking genotype-phenotype correlations emerging between the clinical features and variant type or mode of inheritance (Table 1; Ext Data Fig. 6). Significant differences were found only for movement disorders and febrile seizures. Movement disorders occurred more often in biallelic cases (64%, 21/33) than in dominant ones (10%, 2/20; *P*=1.5 × 10^?4^). Febrile seizures were enriched in dominant cases (41%, 9/22 vs. 3%, 1/36; *P*=0.02), an effect driven entirely by n.4G>A (9/14, 64%), with none (0/8) reported for n.35A>G. Biallelic variants were observed in patients who had not achieved independent sitting (12/37, 32%) compared with 1/20 (5%) of monoallelic cases, and in patients with brain abnormalities (25/41, 61% vs. 8/25, 32%). All six reported deaths (ages 3-19 years; median 10.25 years) occurred in individuals with biallelic variants, due to respiratory, infectious, or epilepsy-related complications. The phenotype was consistent within families, including the severity of ID and the presence of epilepsy or movement disorders. Minor dysmorphic features were commonly observed in patients with available photographs, including a broad forehead, midface hypoplasia, a large open mouth, a small chin, and, in some cases, down-slanting palpebral fissures (Fig. 4b-c).

**Table 1.** Overview of clinical features in patients with monoallelic and biallelic *RNU2-2* variants.

| Group | n.4G>A | n.35A>G | Monoallelic | Biallelic | Total |
| --- | --- | --- | --- | --- | --- |
| Number of patients | 17 | 10 | 27 | 47 | 74 |
| Epilepsy | 16/17 (94%) | 8/10 (80%) | 24/27 (89%) | 41/46 (89%) | 65/73 (89%) |
| *Generalized seizures* | 8/15 (53%) | 6/8 (75%) | 14/23 (61%) | 22/26 (85%) | 36/49 (73%) |
| *Focal seizures* | 10/15 (67%) | 6/8 (75%) | 16/23 (70%) | 9/23 (39%) | 25/46 (54%) |
| *Generalized + focal seizures* | 6/15 (40%) | 4/8 (50%) | 10/23 (43%) | 3/30 (10%) | 13/53 (25%) |
| *Hemicorporal seizures* | 6/14 (43%) | 1/8 (12%) | 7/22 (32%) | 3/21 (14%) | 10/43 (23%) |
| *Tonic-clonic seizures* | 5/14 (36%) | 4/8 (50%) | 9/22 (41%) | 19/26 (73%) | 28/48 (58%) |
| *Clonic seizures* | 6/15 (40%) | 1/8 (12%) | 7/23 (30%) | 7/22 (32%) | 14/45 (31%) |
| *Tonic seizures* | 6/14 (43%) | 2/8 (25%) | 8/22 (36%) | 13/23 (57%) | 21/45 (47%) |
| *Myoclonic seizures* | 4/14 (29%) | 2/8 (25%) | 6/22 (27%) | 20/29 (69%) | 26/51 (51%) |
| *Atonic seizures* | 3/14 (21%) | 3/8 (38%) | 6/22 (27%) | 9/26 (35%) | 15/48 (31%) |
| *Spasms* | 4/14 (29%) | 1/8 (12%) | 5/22 (23%) | 11/28 (39%) | 16/50 (32%) |
| *Non-motor seizures (absences)* | 9/15 (60%) | 1/8 (12%) | 10/23 (43%) | 16/29 (55%) | 26/52 (50%) |
| *Febrile seizures* | **9/14 (64%)** | **0/8 (0%)** | **9/22 (41%)** | **1/36 (3%)** | **10/58 (17%)** |
| *Status epilepticus* | 7/14 (50%) | 6/7 (86%) | 13/21 (62%) | 12/34 (35%) | 25/55 (45%) |
| *Pharmoresistance* | 12/15 (80%) | 4/6 (67%) | 16/21 (76%) | 28/39 (72%) | 44/60 (73%) |
| Hospitalisation | 8/15 (53%) | 5/10 (50%) | 13/25 (52%) | 31/40 (78%) | 44/65 (68%) |
| Feeding issues/ gastrointestinal reflux | 6/15 (40%) | 3/10 (30%) | 9/25 (36%) | 30/45 (67%) | 39/70 (56%) |
| Autism spectrum disorder | 12/17 (71%) | 8/10 (80%) | 20/27 (74%) | 16/38 (42%) | 36/65 (55%) |
| Neonatal findings | 9/16 (56%) | 6/10 (60%) | 15/26 (58%) | 22/46 (48%) | 37/72 (51%) |
| *Neonatal hypotonia* | 7/16 (44%) | 4/10 (40%) | 11/26 (42%) | 16/45 (36%) | 27/71 (38%) |
| *Neonatal feeding problems* | 4/17 (24%) | 4/10 (40%) | 8/27 (30%) | 10/42 (24%) | 18/69 (26%) |
| Brain MRI anomaly | 5/16 (31%) | 3/9 (33%) | 8/25 (32%) | 25/41 (61%) | 33/66 (50%) |
| Sleep disorders | 9/16 (56%) | 1/10 (10%) | 10/26 (38%) | 20/40 (50%) | 30/66 (45%) |
| Movement disorder | **2/13 (15%)** | **0/7 (0%)** | **2/20 (10%)** | **21/33 (64%)** | **23/53 (43%)** |
| Constipation | 7/15 (47%) | 3/10 (30%) | 10/25 (40%) | 18/40 (45%) | 28/65 (43%) |
| Dysmorphic features | 8/16 (50%) | 5/10 (50%) | 13/26 (50%) | 16/43 (37%) | 29/69 (42%) |
| Hypersialorrhea/drooling | 8/15 (53%) | 3/10 (30%) | 11/25 (44%) | 15/40 (38%) | 26/65 (40%) |
| Eyes/vision abnormalities | 3/15 (20%) | 3/10 (30%) | 6/25 (24%) | 16/41 (39%) | 22/66 (33%) |
| Bone/skeletal anomalies | 4/15 (27%) | 2/10 (20%) | 6/25 (24%) | 15/43 (35%) | 21/68 (31%) |
| Ataxia | 5/15 (33%) | 5/8 (62%) | 10/23 (43%) | 7/34 (21%) | 17/57 (30%) |
| Short stature | 6/17 (35%) | 5/10 (50%) | 11/27 (41%) | 8/39 (21%) | 19/66 (29%) |
| Joint hyperlaxity | 4/15 (27%) | 2/10 (20%) | 6/25 (24%) | 13/40 (32%) | 19/65 (29%) |
| Microcephaly | 2/16 (12%) | 2/10 (20%) | 4/26 (15%) | 13/40 (32%) | 17/66 (26%) |
| Failure to thrive / growth retardation | 2/16 (12%) | 2/10 (20%) | 4/26 (15%) | 13/41 (32%) | 17/67 (25%) |
| Prenatal findings | 1/16 (6%) | 1/10 (10%) | 2/26 (8%) | 6/43 (14%) | 8/69 (12%) |

MRI: magnetic resonance imaging. Clinical features are listed by frequency across the entire patient cohort, with significant differences between monoallelic and biallelic groups shown in bold.

**Fig 6.**
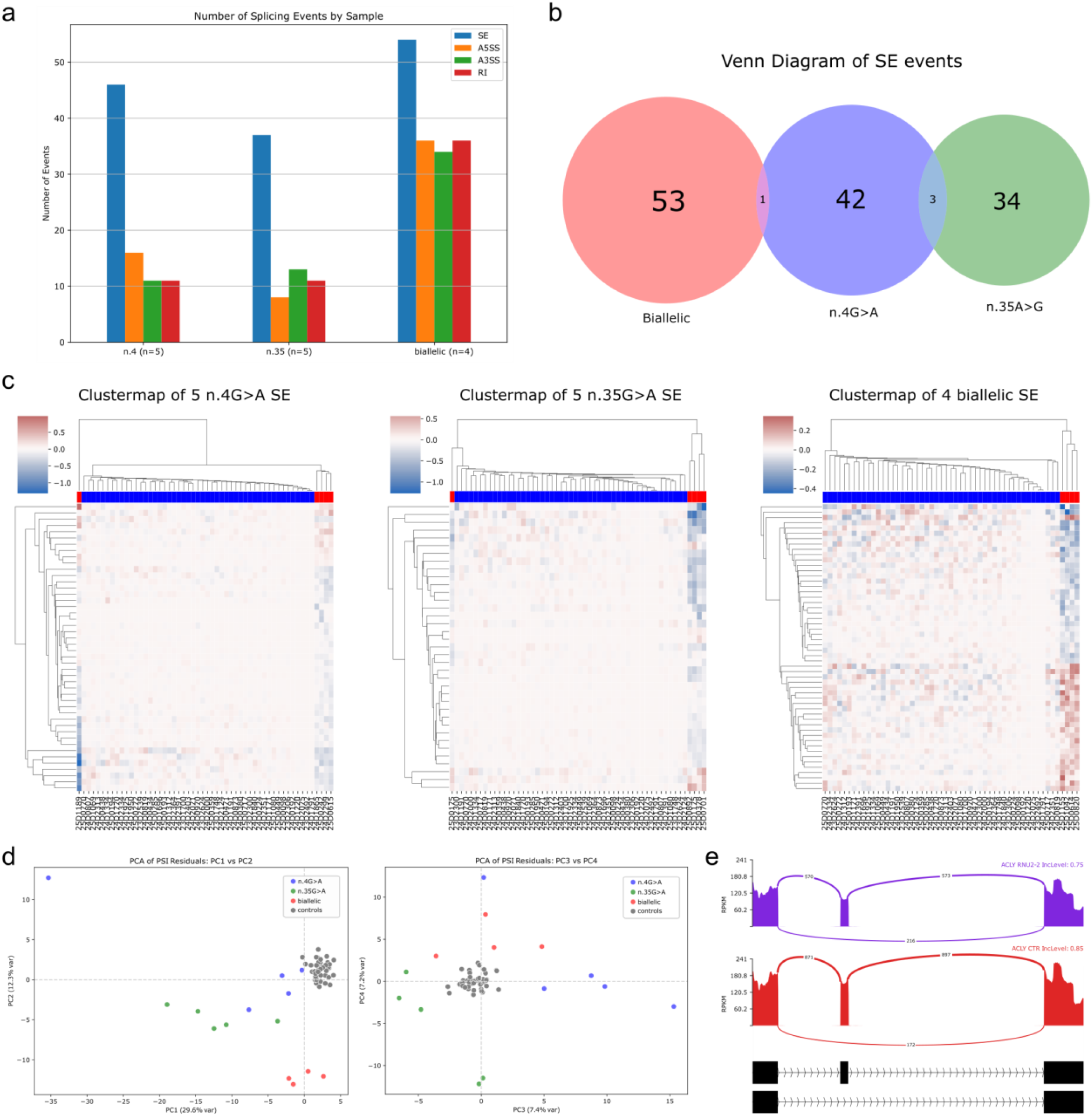
Variant-specific alternative splicing perturbations in *RNU2-2* variant carriers. **a**, Number of significant alternative splicing events (ΔPSI > 0.05) detected using rMATS and linear regression (p-val<0.01), comparing *RNU2-2* individuals (dominant: n.4G>A, *n*=5; n.35A>G, *n*=5; recessive: *n*=4) to 49 controls. Splicing categories are colour-coded: exon skipping (SE, blue), alternative 5′ splice sites (A5SS, orange), alternative 3′ splice sites (A3SS, green), and intron retention (RI, red). **b**, Venn diagram showing minimal overlap of exon skipping events shared among dominant (n.4G>A, n.35A>G) and recessive (biallelic) variant groups. **c**, Clustermaps of alternative splicing events for individuals carrying n.4G>A (left, *n*=5), n.35A>G (middle, *n*=5), and biallelic variants (right, *n*=4). **d**, Principal component analysis based on residuals of PSI values after linear regression showing separation of controls (grey, *n*=49) from individuals with n.4G>A (*n*=5, blue), n.35A>G (*n*=5, green), and biallelic variants (*n*=4, red). Right panel: PC1 vs. PC2; left panel: PC3 vs. PC4. **e**, Sashimi plots illustrating an exon-skipping isoform shift in *ACLY* observed in *RNU2-2* variant carriers compared to controls.

The five patients with monoallelic *de novo* variants other than n.4G>A and n.35A>G showed even more variable clinical presentations. One patient died at 6 days of life from status epilepticus; the four other patients had mild ID without epilepsy (*n*=3) or moderate ID with epilepsy (*n*=1).

### Predicted impact of *RNU2-2* variants

To predict the impact of *RNU2-2* variants, we considered their distribution across U2 structural and functional domains. U2-2 can be divided into three functional domains, each remodelled to different extents throughout the splicing cycle and snRNP biogenesis. The 5’ domain forms four partially mutually exclusive structures: (1) the intramolecular branch-point-interacting stem loop (BSL; nt 25– 45)^26^ and (2) the stem-loop I (SLI; nt 7–26)^27^, (3) the intermolecular U2/U6 helix II (nt 1-13)^1^, and (4) the U2/U6 helix Ia and Ib (nt 20-28)^28^. The branch interacting region oscillates between BSL conformation and stable intron binding via formation of U2/BS branch helix (nt 32-44)^1,29^. The 3’ end domain encompasses five structural elements: stem-loops IIa (SLIIa; nt 47-66)^30^ and IIb (SLIIb; nt 68-84)^26^, the Sm binding site (nt 98-107)^31^, and 3’ stem-loops III and IV (SLIII; nt 112-144/SLIV; nt 147-184)^32,33^. Identified variants were mapped onto published U2 snRNP or spliceosome structures^34,35^ to assess potential structural impact.

*RNU2-2* variants at the 5’ end likely destabilise U2/U6 helix II by disrupting key Watson-Crick base pairs:^27^ n.4G>A breaks the G4–C94 pair, n.5C>A disrupts C5–G93, and n.6T>C alters U6–A92 (Fig. 5). These changes may impair tri-snRNP recruitment to the pre-spliceosome and reduce splicing efficiency.^36-38^ n.7_8insA, n.20G>A, n.23A>C, n.24A>G and n.26A>C would affect SL1 stability and directly or indirectly alter U2/U6 helix II formation. Additionally, n.20G>A, n.26A>C, n.27T>A, and n.28C>G may destabilize helices Ia and Ib by replacing Watson-Crick base pairs with noncanonical pairs, perturbing active site formation.

Some variants are also found within the BSL and likely affect its stability. For instance, n.40C>T, by creating a A30–U40 base pair, would hyper-stabilise the BSL, which was previously linked to reduced splicing fidelity in yeast^26^. Likewise, n.27T>A and n.40C>G may stabilize the BSL by creating A-Ψ and G-A base pairs replacing U-Ψ and C-A mismatches.^39^ Conversely, n.26A>C, n.28C>G, n.44T>C, and n.45C>T may disrupt BSL integrity, potentially perturbing Prp5 action.^40^ Interestingly, n.35A>G alters U2’s invariant A35 that pairs with the conserved U upstream of the BS, which may increase pairing flexibility, promoting cryptic branch site usage^41^ (Fig. 5). Of note, n.28C>G, which may affect both BSL formation and the active site, is a recurrent somatic variant detected in cancers.^21^

In contrast, variants in SLIIa, the Sm site, and the 3′ stem loops SLIII/SLIV likely impair U2 snRNP biogenesis and nuclear import, destabilizing U2 and preventing proper spliceosome assembly. These changes may effectively behave as null alleles like gene deletions, as the affected U2 molecules may fail to reach functional spliceosomes.

### Impact of *RNU2-2* variants on lymphocyte transcriptome

We previously showed that pathogenic *RNU4-2* variants lead to splicing defects primarily affecting alternative 5′ splice sites (5’SS), detectable in cultured lymphocytes ^16^. Here, we investigated splicing alterations in 14 individuals with either dominant (n.4G>A, *n*=5; n.35A>G, *n*=5) or recessive (*n*=4) *RNU2-2* variants and compared them with data of 49 patients with other disorders (controls). This first approach did not reveal a consistent transcriptomic signature shared by *RNU2-2* variant carriers. To increase the specificity of our analysis, we applied a linear regression on percent-spliced-in (PSI) values (ΔPSI > 0.05), incorporating age, gender, and cell composition as covariates. This approach effectively increased the separation of cases and controls on PCA and predominantly detected exon skipping (SE) events in dominant (n.4 and n.35) cases, whereas recessive cases showed a broader spectrum, affecting all splicing categories (5’SS; 3’ splice sites, 3’SS; and intron retention, RI) (Fig. 6a). Nevertheless, exon skipping events showed minimal overlap between all three groups (Fig. 6b). Clustermaps showed predominantly increased exon skipping in n.4 and n.35, while recessive cases exhibited both increases and decreases (Fig. 6c). All aberrant events were also observed in controls (Fig. 6d-e), indicating that *RNU2-2* variants only subtly perturb alternative splicing rather than introducing novel junctions in lymphocytes.

### Impact of *RNU2-2* variants on blood methylome

We next analysed DNA methylation profiles in 24 individuals with *RNU2-2* variants (8 with n.4G>A, 6 with n.35A>G, and 10 with biallelic variants) and compared them with 68 controls to identify potential episignatures, using a similar methodology to *RNU4-2*^16^. Cases and controls were matched for age at sampling, and analyses also corrected for age, sex, and cell composition. Consistent with the transcriptomic findings, patient groups appeared heterogeneous, limiting statistical power for separate variant-specific analyses. We then implemented a combined model simultaneously evaluating all three variant types while allowing for both shared and variant-specific effects. Overall, methylation signatures associated with *RNU2-2* variants were subtle, heterogeneous, and weaker than previously reported for *RNU4-2* (Fig. 7a). PCA across all 201 differentially methylated positions (DMPs) showed that n.4G>A carriers exhibited the strongest signal, although only ∼25% of variance was captured by PC1 (Ext Data Fig. 7a; Supplementary Table 5). Separation of n.35A>G and biallelic carriers from controls became apparent only at higher PCs, reflecting the minor effect size. Excluding n.4G>A carriers improved visualization, with n.35A>G and biallelic variants partially separating from controls along PC2 (Ext Data Fig. 7b-c). Methylation profiles showed that n.4G>A carriers were largely hypomethylated, whereas n.35A>G carriers displayed modest hypermethylation (∼2.5% Δβ), and biallelic carriers had heterogeneous patterns without consistent hypo-or hypermethylation. Cross-validation with a single four-class SVM classifier confirmed the strongest signature for n.4G>A (sensitivity 100%), with lower performance for n.35A>G (83%) and biallelic variants (80%), and an overall specificity of 87% (Fig. 7b; Ext Data Fig. 7d). Overall, methylation differences between variant carriers and controls were less than 5% Δβ, near the technical detection limit, suggesting subtle and heterogeneous effects across variants

**Fig 7.**
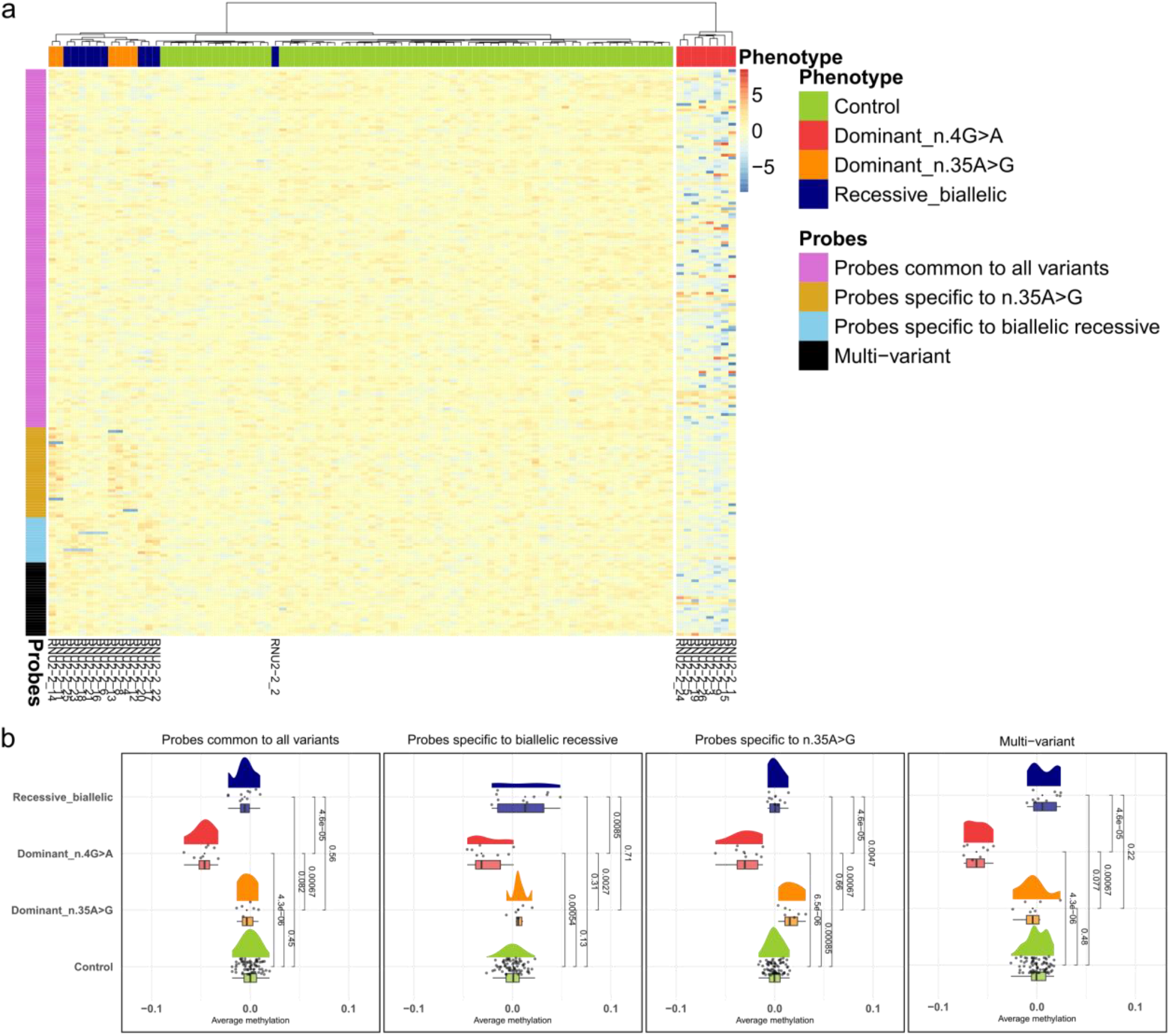
Methylation profiles across differentially methylated probes in *RNU2-2* variant carriers and normal controls. In all panels, controls are shown in green, dominant n.4G>A carriers in red, dominant n.35A>G carriers in orange, and recessive biallelic cases in blue. **A**, Heatmap of adjusted methylation levels at differentially methylated CpGs. Selected probes are represented in rows while patients and normal controls are represented in columns after hierarchical clustering of their methylation profiles. Probes are grouped according to their association pattern: probes common to all variants, probes specific to n.35A>G, probes specific to biallelic recessive variants, and probes specific to n.4G>A. Samples are annotated by type (Control, Dominant n.4G>A, Dominant n.35A>G, Recessive biallelic) but sample clustering was unsupervised and blind to this classification. **b**, Raincloud plots showing the average methylation level per sample within each probe association pattern. Normal controls and variant carriers display significantly different distributions, with p-values from two-sided Wilcoxon rank-sum tests indicated above the plots.

## Discussion

Although historically overlooked due to their high conservation and redundancy, snRNA genes are now recognised as major contributors to genetic disorders. In this study, we analysed all annotated snRNA genes, hypothesising that some ‘pseudogenes’ may in fact be functional disease genes. Our main finding was the identification of biallelic variants in *RNU2-2* as the cause of a recessive NDD. Clinically, this recessive NDD closely resembles the recently described dominant *RNU2-2*–associated disorder caused by n.4G>A and n.35 A>G,^17,20^ but appears at least twice more common. The clinical phenotype associated with dominant pathogenic or biallelic *RNU2-2* variants is characterized by neurodevelopmental delay and predominantly severe ID, with epilepsy occurring in 86% of cases, typically with onset before the age of 3 years in 80%. Movement disorders are more frequent in patients with biallelic variants. Excluding single *de novo* variants other than n.4G>A and n.35A>G, we estimate that dominant and recessive *RNU2-2* variants altogether account for 0.35% (63/∼18,000; PFMG cohort) of NDDs, which is close to the prevalence of ReNU syndrome^14,16^. While some biallelic variants may be non-pathogenic, the consistent clinical phenotypes and predicted structural impact support a likely disease contribution for most variants. Although we applied strict selection criteria, relaxing these revealed additional families, suggesting that the true prevalence of this disorder may even be underestimated. Compared to *RNU4-2, RNU2-2* shows lower constraint and greater tolerance to variation,^14^ aligning with its association predominantly with recessive disorders, in contrast to the dominant ReNU syndrome caused by *RNU4-2* variants.

*RNU2-2* exhibits a complex spectrum of variant effects that challenge simple dominant versus recessive classification. Our results show that *de novo* variants in snRNA genes can underlie recessive as well as dominant inheritance, reflecting the high mutability of functional snRNA genes.^22^ Therefore, *de novo* variants present at low frequency in population databases should not be dismissed and a second pathogenic variant in *trans* should always be searched for. Interpreting *de novo* variants beyond n.4G>A and n.35A>G when no second pathogenic allele is present remains challenging. Several of these variants were found in *trans* with another allele, supporting a recessive mode of inheritance and suggesting they may not be pathogenic on their own. This appears to be the case for many recurrent, low-frequency variants, such as n.45C>T and those in the 3′ domain, which are typically inherited and likely require biallelic occurrence to cause disease. In contrast, other *de novo* variants near n.4 and n.35 are extremely rare or absent from population databases, recur *de novo* across unrelated individuals, and are under stronger negative selection. These variants, observed either alone or in *trans* with others, are consistently associated with phenotypes compatible with *RNU2-2*-NDD. These observations support a gradient-of-impact model, in which certain heterozygous variants may contribute to dominant disease with variable penetrance, while they can also cause a classic recessive phenotype when associated with a second pathogenic allele in *trans*.

These observations must be interpreted in light of predicted variant effects on U2 snRNA and the presence of additional U2 copies, including canonical *RNU2-1* paralogs. U2 largely contributes to defining splicing outcomes by selecting the intronic BS (‘YNYURAY’ consensus in metazoans) and thereby the acceptor splice sites.^42^ Prior to its integration within the spliceosome, U2 snRNAs undergo a maturation process that includes Sm-core assembly, nuclear reimport, and protein recruitment to form the U2 snRNP.^32,33,43-46^ The 3′ domain mainly mediates structural interactions necessary for U2 snRNP biogenesis and spliceosome assembly^32,46,47^. For example, SLIIa binds the central U2 snRNP subunit SF3B1.^43,48^ In contrast, the 5′ domain plays a more complex role via highly conserved RNA/RNA interactions contributing to spliceosome assembly, active site formation, and branch point recognition. During early spliceosome assembly, U2 initially probes the branch site (BS) and then forms a stable U2/BS branch helix.^26^ This process requires the PRP5 ATPase, which remodels the BSL and enforces fidelity of BS recognition.^36^ Formation of the branch helix may trigger the formation of SLI.^48^The latter is subsequently disrupted to allow tri-snRNP recruitment via creation of the U2/U6 helix II, forming a precatalytic spliceosome. Upon spliceosome activation, U2 forms two additional contiguous helices with U6 (Ia, Ib) that, along with helix II, flank and stabilise the RNA-based active site.^49,50^ Our predictions suggest that variants in 5′ region of U2-2, especially within the U2/U6 helix II and branch site recognition sequence, have the strongest impact. Two recurrent variants (n.4G>A, n.35A>G) act dominantly, likely incorporating into spliceosomes and causing exon-skipping defects in heterozygotes. Nearby variants may have similar but milder effects, resulting in reduced penetrance in heterozygotes and full penetrance when combined with a second pathogenic allele. In contrast, variants in the Sm site or 3′ stem loops may prevent U2 snRNP maturation and spliceosome incorporation, acting as null alleles analogous to larger indels and whole gene deletions. These null alleles are tolerated when heterozygous but pathogenic in homozygous or compound heterozygous states, suggesting an essential role of *RNU2-2* in human brain development. Overall, pathogenic *RNU2-2* variants likely disrupt splicing and drive NDDs through diverse mechanisms, ranging from weakening U2/U6 interactions, impairing branch site recognition to blocking snRNP biogenesis, resulting in a spectrum of splicing defects.

Contrary to *RNU4-2, RNU2-2* variants exert a subtler effect on splicing in blood, primarily causing exon skipping for dominant variants and broader splicing alterations for recessive variants. These findings align with the lower expression of *RNU2-2* than *RNU2-1* in blood, which contrasts with its higher relative expression in the brain,^17,20^ and is consistent with the primarily neurodevelopmental features of *RNU2-2*–associated disorders. Similarly, DNA methylation analyses in *RNU2-2* variant carriers revealed only modest, variant-specific changes. These results should be interpreted cautiously and validated in larger cohorts, as some differences may reflect stochastic variability rather than consistent, biologically meaningful episignatures, and may also mirror the variants’ diverse functional effects.

Despite the size of the PFMG cohort (>26,000 individuals), we did not identify additional significant snRNA-disease associations. This likely reflects limited statistical power to detect variants in snRNA genes causing rarer disorders, as known disease associations with *de novo* variants in *RNU5B-1*,^16,17^ or recessive variants in *RNU4-2, RNU6ATAC*^51^ and *RNU12*^12,13^ did not reach significance. Increasing statistical power will require pooling international cohorts for robust genotype-phenotype association studies. Another important limitation of our study is the reliance on short-read genome sequencing, which cannot accurately resolve highly similar snRNA loci, including most U1 and U2 canonical genes, due to the presence of (nearly) identical copies contained in regions spanning >5–10 kb in sequence homology. The analysis of these regions thus requires long-read sequencing technologies.

In conclusion, our results support a continuum of dominant and recessive NDDs caused by *RNU2-2* variants, which altogether are nearly as prevalent as ReNU syndrome. They also underscore the challenges of interpreting variants, especially those occurring d*e novo*, in these highly mutable loci, and suggest that additional, rarer snRNA-associated disorders remain to be discovered.

## Acknowledgements

We are deeply grateful to the patients and their families for their participation in this study. Patients included in this study were diagnosed through multiple studies. Patients from the PFMFG received funding from Plan France Médecine Génomique 2025 (PFMG2025). Cases identified by the Broad Center for Mendelian Genomics (A.O’.D-L. and S.L.S.) were supported by the National Human Genome Research Institute (NHGRI) grants UM1HG008900 and U01HG011755. Genetic analyses (cases from Gleeson lab) were supported by the Gabriella Miller Kids First Pediatric Research Program, funded by the Common Fund of the NIH Office of the Director, with sequencing performed at the Broad Institute Sequencing Center (U24HD090743). Genetic analyses of Australian cases were supported by the Genetic Basis of Epilepsy Research program (NHMRC grants GNT1172897, GNT2033247, GNT2006841) and by Victorian State Government Operational Infrastructure Support and the NHMRC IRIISS. M.F.B., M.S.H., and I.E.S. were supported by an Australian Medical Research Future Fund Genomics Health Futures Mission Grant (2007707). Part of the results has been supported by the RNU-SPLICE project, financed by Health philantropic program of Mutuelles AXA dedicated to supporting innovative research projects in France (to C.N.). The structural interpretation was conducted by C. Charenton. and M.A. as part of the ERC Starting Grant project " SPLIFEM ". A.S. received a grant from Region Normandie and GIRCI Nord Ouest (FHU-A2M2P). This study uses resources generated by the ENCODE Consortium, 1) the ENCODE Data Analysis Center (ZLab at UMass Medical Center), Henry Pratt, Jill Moore, Michael Purcaro, and Zhiping Weng for providing ENCODE cCREs data, 2) Bernstein Lab at the Broad Institute for the H3K27Ac peaks in H1-hESC, 3) Thomas Gingeras lab at Cold Spring Harbor Laboratory for generating the small RNA-seq data from six human embryonic brain regions.

N.W. is supported by a Wellcome Career Development Award (grant no. 305292/Z/23/Z) and a research prize from the Lister Institute. HCM is funded by ALSAC of St. Jude Children’s Research Hospital. K.Õ. was supported by Estonian Research Council grants PRG471 and PRG2040. Health Research Council of New Zealand and Curekids New Zealand supported the recruitment, phenotyping and sequencing of the New Zealand participants. A.K., F.J.K., D.W., L.F., and P.S-W. are members of the European Reference Network for Developmental Anomalies and Intellectual Disability (ERN-ITHACA). Support for title page creation and format was provided by AuthorArranger, a tool developed at the National Cancer Institute.

## Contributions

E.L. performed the *in silico* analyses of snRNA genes, burden tests, genotype-phenotype correlations, and statistical analyses. G.L., C.N., V.B., P.M., and J.T. contributed snRNA variant data from the PFMG cohort. B.C. and T.B. performed RNA sequencing and data analysis. A.S. and C.C. carried out DNA methylation experiments and analysed episignatures. M.A. and C.C. conducted the structural analyses. G.L., C.N., and C.L. collected clinical information. C.N., G.L., C.C., A.S., and B.C. contributed to sections of the manuscript, and C. D. drafted the article. C. D., C.N., and G.L. supervised the study. All other authors contributed molecular and/or clinical data, provided intellectual input, and approved the final manuscript.

## Data availability

Variant details will be submitted to ClinVar for publication. RNA-seq and methylation data will be deposited in the European Genome-phenome Archive (EGA, http://www.ebi.ac.uk/ega), hosted by the EBI. Both datasets will be subject to a Data Processing Agreement, and access requests will be reviewed by a Data Access Committee to ensure compliance with ethical and legal standards. Due to ethical considerations, individual genome data cannot be made publicly available. Controlled access is required to safeguard participant privacy and to comply with data protection regulations, including the GDPR in Europe. Access to genome data from the PFMG2025 cohort is governed by French data protection laws and is only possible via the Collecteur Analyseur de Données (CAD). More details can be found on the PFMG2025 website: https://pfmg2025.fr/le-plan/collecteur-analyseur-de-donnees-cad/.The coordinates of the ENCODE Registry of candidate cis-Regulatory Elements (cCREs) in the human genome^23^ and bigwig files concerning the peaks of histone H3 acetylation of lysine 27 (H3K27Ac) obtained for the H1 human embryonic stem cell line (H1-hESC)^24^ were downloaded through the UCSC Table Browser^52^. Bigwig files concerning small RNA-seq data from six human embryonic brain regions were downloaded from the ENCODE portal^53^ (http://www.encodeproject.org/) with the following identifiers: ENCFF013RLG, ENCFF029RIV, ENCFF034QAV, ENCFF197SSE, ENCFF221KEN, ENCFF343ZBS, ENCFF405QIN, ENCFF532SOY, ENCFF654ONK, ENCFF738LDD, ENCFF870FMA, ENCFF887TOS, ENCFF106ESQ, ENCFF222WBQ, ENCFF250WEA, ENCFF254UEQ, ENCFF319GRF, ENCFF425WUZ, ENCFF443ONL, ENCFF820JTT, ENCFF897IWP, ENCFF915WAC, ENCFF946YVE and ENCFF965GHD.

## Code availability

No custom code was developed for this study. All analyses were performed using existing, publicly available pipelines and software tools listed in the methods and reporting summary.

## Methods

### Inclusion & Ethics statement

This study was conducted in accordance with the ethical standards and regulations of all participating countries. Written informed consent was obtained for all patients from their parents or legal guardians, with an additional consent form for families agreeing to the publication of photographs. For genetic analyses, patient samples were pseudonymized at each participating center. Information on the patients’ sex (but not gender) was extracted from clinical records. The promoters of this research study are Assistance Publique–Hôpitaux de Paris (AP-HP) for hospitals associated with the SeqOIA laboratory (project ID APHP241333) and Grenoble-Alpes University Hospital (CHU Grenoble-Alpes, research ID 19814188) for hospitals affiliated with the Auragen laboratory. Ethical approval was obtained from the University Hospital Essen (24-12010-BO) and the Comité Éthique et Scientifique pour les Recherches, les Études et les Évaluations dans le domaine de la Santé (CESREES; reference 21082803 Bis / 2038764). AP-HP has received an authorization from the Commission Nationale de l’Informatique et des Libertés (CNIL; reference HGTHGT/MFIMFI/AR2426865; request no. 924924336666) for data processing. Additional approvals were obtained from the ethics committee of CHU de Nantes (CCTIRS number 14.556) and from CPP Ouest V (File 06/15, Ref MESR DC 2017 2987; approval date 04/08/2015). For methylation analyses, DNA from patients and controls had been previously collected in a medical context for genetic testing, with written consent including authorization for research use of leftover material. Control samples consisted of individuals without neurodevelopmental disorders, either unaffected relatives or persons tested presymptomatically for other conditions who were found not to carry pathogenic variants. DNA samples used for methylation profiling were stored within the genetics biobank of the CRBi, Rouen, France (collection DC 2008-711, authorization MCRBi/2024/02). The use of these samples was approved by the CERDE ethics committee of Rouen University Hospital (notification E2023-13). Researchers and clinicians from all contributing centers participated throughout the study, from design and implementation to data collection, analysis and manuscript preparation, and are listed as coauthors of this article.

### snRNA genes

Genes and pseudogenes coding for small nuclear RNAs (snRNAs) were retrieved from Ensembl 113 BioMart^54,55^ by filtering on gene type ‘snRNA’. Of the 2,094 genes, we excluded the ones placed in contigs, scaffolds or patches, keeping 1,901 genomic regions located in regular chromosomes. Of these, 1,741 genes encode snRNAs that are components of the spliceosome. We downloaded through the UCSC Table Browser^52^: the coordinates of the ENCODE Registry of candidate cis-Regulatory Elements (cCREs) in the human genome^23^ and 2) the peaks of histone H3 acetylation of lysine 27 (H3K27Ac) obtained for the H1 human embryonic stem cell line (H1-hESC)^24^. We annotated the 1,901 snRNAs by analysing their genomic coordinates for overlaps with ENCODE cCREs and H3K27Ac peaks. The RNU genes were further annotated for their hypermutability.^22^ We kept for further analyses 200 putative functional snRNAs that either overlap with ENCODE cCREs, are considered hypermutable, or are approved by the HUGO Gene Nomenclature Committee (HGNC).

### Small RNA-seq data from embryonic brain tissues

We analysed the expression level of the 1,901 snRNAs using small RNA-seq data generated from six human embryonic brain regions by the ENCODE Consortium^56^: diencephalon (GSE78292), temporal lobe (GSE78303), occipital lobe (GSE78298), frontal cortex (GSE78293), parietal lobe (GSE78299) and cerebellum (GSE78291). We downloaded 24 bigwig files from the ENCODE portal^23^ containing signals for ‘all reads’ and ‘unique reads’, for plus and minus strands, from the default anisogenic replicate. For each snRNA genomic coordinates, we calculated the maximum normalized RNA-seq signal obtained from the bigwig files for the corresponding gene strand, as well as the number of covered bases. For plotting, we considered only snRNAs for which at least 50% of the expected number of bases for the specific snRNA type were covered in at least one tissue when considering ‘all reads’. The expected number of bases for each snRNA type were 164 (U1), 191 (U2), 141 (U4), 116 (U5), 107 (U6), 63 (U7), 127 (U4ATAC), 126 (U6ATAC), 134 (U11), 150 (U12) and 40 (other snRNAs).

### Patient cohorts

We first investigated monoallelic and biallelic variants in *RNU2-2* in patients who underwent genome sequencing as part of the diagnostic process in France via Plan France Médecine Génomique 2025 (PFMG2025)^25^ on one of the two national clinical sequencing laboratories, SeqOIA (https://laboratoire-seqoia.fr/) and Auragen (http://www.auragen.fr/) between 2019 and May 2025. Data were generated in two subsets (in SeqOIA and Auragen laboratories) and analysed separately and combined, as bioinformatics pipelines differed slightly. At the time of the study, the cohort included 26,911 patients with rare disorders (15,945 from SeqOIA and 10,966 from Auragen). For monoallelic variants, the inclusion criteria were: 1) a *de novo* variant observed in fewer than 50 heterozygotes in the *All of Us* database, or 2) a previously reported pathogenic monoallelic variant (n.4G>A or n.35A>G) with unknown inheritance. Applying these criteria, 32 unrelated patients were included in the study.

For biallelic variants, we initially considered cases presenting either (1) a homozygous variant, or (2) a compound configuration in which one variant was inherited from a parent and the second was either inherited from the other parent or occurred *de novo* in *trans* with the inherited variant (when both parents were sequenced), or was absent in the sequenced parent (when only one parent was sequenced). Candidate variants had to be present in fewer than three homozygotes in gnomAD_v3 and in fewer than five homozygotes in our internal databases. This first analysis identified 88 patients from 55 families in SeqOIA and 33 families in Auragen. As several of these variants appeared too frequent to be consistent with monogenic inheritance, we refined our inclusion criteria to: (1) biallelic variants observed in fewer than 200 heterozygotes and absent in the homozygous state in the *All of Us* database, or (2) biallelic variants in which one allele (possibly more frequent) occurred in *trans* with a recurrent variant (identified in at least three independent recessive cases). For *RNU2-2* variant frequency estimation, we transitioned from gnomAD_v3 to the *All of Us* database, as it encompasses a larger cohort of genomes from healthy individuals, thereby providing greater power to assess population-level variation. Based on this refined strategy, 47 families were retained for analysis. All variants were reviewed using IGV; in particular, for biallelic variants, phasing in *trans* (compound heterozygosity) was assessed in IGV when parental samples were not available. Variant nomenclature was systematically checked with Variant Validator (https://variantvalidator.org/service/validate/).

In addition, we collected data on 26 further patients carrying monoallelic or biallelic *RNU2-2* variants, identified either in diagnostic or research contexts through national networks or established collaborations. This cohort comprised 10 patients with monoallelic variants (n.4G>A, *n*=7; n.35A>G, *n*=2; n.38A>G, *n*=1) and 16 patients with biallelic variants, originating from the US (*n*=6), France (*n*=5), Germany (*n*=4), New Zealand (*n*=3), Egypt (*n*=3), Australia (*n*=2), Denmark (*n*=1), Turkey (*n*=1) and Brazil (*n*=1). Nineteen patients had prior genome sequencing, while in seven cases the variants were identified by Sanger sequencing. None of these patients had been previously published, and we carefully verified the absence of duplicates among individuals carrying the same variant by cross-checking year of birth and initials.

### Variant classification

We classified variants according to the ACMG/AMP criteria using recommendations of Ellingford *et al*.^57^ For *de novo* variants, PS2 was applied but downgraded to PS2_Supporting in light of the hypermutable nature of the gene. PM2_Supporting was applied to variants absent or very rare in gnomAD v3 (allele count <10) and *All of Us* (allele count <20). PM1 was applied to variants located in functional domains (n.4 - n.45: U2/U6 helix II, Stem loop I, and BSL; and n.99 - n.104: Sm site).

PM1_Supporting was applied to variants in n.48-n.66 (Stem loop IIa), n.128 - n.132 (Stem loop III), and n.142 - n.183 (Stem loop IV). PS4_Supporting was applied for variants present in two unrelated patients, PS4_Moderate when observed in three to four unrelated patients, and PS4 when present in five or more unrelated patients. PP1 was applied if a variant co-segregated with disease in two or three affected family members, PP1_Moderate was applied when co-segregation was observed in five family members (affected or unaffected) or across multiple unrelated families. PM3 was assigned to variants detected in *trans* with a likely pathogenic or pathogenic variant. Finally, PP5_Moderate was applied to variants previously reported as pathogenic (n.4G>A and n.35A>G).

We also annotated variants with MobiDeep available on Mobidetails website (https://mobidetails.chu-montpellier.fr).^58^ MobiDeep is a metascore for non-coding variants based on a multilayer perceptron using 5 features: REMM v.0.4, CADD 1.7, GPN-MSA, and two conservation scores (Cactus 241-way vertebrates and PhyloP primates) to capture different evolutionary depths. Variants were stratified according to MobiDeep thresholds: <0.6 (neutral), >0.6 (likely deleterious), and >0.9684 (high-confidence deleterious).

### Sanger sequencing

Sanger targeted sequencing was performed to screen for variants in *RNU2-2* and/or to perform segregation analysis. PCR amplification of *RNU2-2* was performed using the HotStarTaq Master Mix Kit (Qiagen, Ref: 203445) with the following primers: F: 5’-CAAACACGCGTCATTCAACACAC-3’; R: 5’-ATAACTGGTTGGAAGATGGGAAGG-3’, according to the manufacturer’s instructions. Forward and reverse sequencing reactions were performed using the BrilliantDye Terminator v1.1 Cycle Sequencing Kit (Nimagen; Ref: BRD1-1000) or the BigDye Terminator v3.1 Sequencing Kit (Life Technologies, Ref: 4337457). ExoSAP-Purified sequencing products (ExoSAP-IT, Applied Biosystem, Ref: 78205) were run on Pop-7 polymer (Life Technologies, Ref: 4335615) using an ABI 3730 or 3730XL automated sequencer (Applied Biosystems). Sequences were analysed using Geneious Prime® 2019 (Biomatters Ltd.) or Seqscape v2.6 software (Applied Biosystems).

### Clinical analyses

Clinical data were retrospectively collected from the referring physician using an anonymized Excel sheet, as previously published^16^. We collected categorical data on 63 clinical features from 66 patients. For clustering and statistical comparisons, we used 58 features, except in the analysis comparing variants n.4G>A and n.35A>G, where 54 features were included since the remaining features showed no variation among patients. Data were converted to a 0-1 scale, with 0 representing a more favourable phenotype presentation and 1 a more severe phenotype. Hierarchical clustering was performed using pheatmap R package, performing Z-score scaling for each row (across different patients), and ward.D2 clustering method keeping missing values. PCA was generated after replacing missing data by 0 and performing variable scaling. Microcephaly was defined as head circumference (HC) measurements < 3rd percentile. We used charts established by Fenton et al^59^ to calculate HC percentile at birth and define congenital microcephaly. Fisher’s tests (two-sided; 2×2, 2×3 or 2×4 contingency tables) adjusted for multiple comparisons using Bonferroni correction were used to compare clinical features in different U2 domains (n.4G>A, n.35A>G, other *de novo* variants vs. biallelic variants) for 63 clinical features.

### Prediction of variant impact

Structural analysis of variants and corresponding figures were performed using the PyMol v3.0.0 visualisation software^60^ on published coordinates of the human U2 snRNP structures: PDB 7Q3L^34^ for the BSL structure and 5XJC^35^ for the representation of the rest.

### RNA sequencing

RNAseq experiments were conducted following a similar protocol and workflow previously described for *RNU4-2*^*16*^. Briefly, PBMCs were isolated from 2-4 mL of EDTA blood and cultured in lymphocyte-stimulating medium for 48–72 h. RNA was extracted and stranded RNA-Seq libraries prepared from 100 ng total RNA using the SureSelect XT-HS2 kit (Human All Exon V8 capture probes; Ref: G9774C), followed by sequencing on an Illumina NextSeq 550 to obtain ∼25–30 million paired-end reads per sample. Reads were aligned to GRCh38 with STAR v2.7.11a. Quality control was performed with FastQC v0.11.3 and Fastp v0.23.4. Cell type composition was estimated using CIBERSORTx v1.0 with the LM22 signature matrix. For splicing analysis, we focused on 14 individuals carrying either dominant (*n*.4G>A, *n*=5; *n*.35A>G, *n*=5) or recessive (*n*=4) *RNU2-2* variants that we compared to 49 controls (probands without *RNU2-2* variants). rMATS-turbo v4.3.0 was used to detect alternative splicing events with the following parameters: `-t paired –anchorLength 1 –libType fr-firststrand –novelSS –variable-read-length –allow-clipping`. rMATS outputs were filtered for mean coverage >10 and ΔPSI >0.05. For each splicing event, we fitted a linear regression model adjusting for age, sex, and estimated cell composition using Ordinary Least Squares with statsmodels (v.0.14.5) in python (v.3.12.2). P-values were obtained for association with affected status; significant splicing events were defined as those with p-value < 0.01, retaining only the most significant event per gene. To visualize results, the linear regression model was trained on the 49 control samples and applied to both controls and affected individuals; residual PSI values were then used for principal component analysis (PCA) and clustermaps for the splicing categories: exon skipping (SE), 5’ and 3’ splice sites (5’SS, 3’SS), and intron retention (RI).Sashimi plot was performed with rmats2sashimi.

### DNA methylation study

DNA methylation analysis was also conducted as previously described.^16^ Genomic DNA was extracted from whole blood and subjected to bisulfite conversion. DNA methylation profile was then derived using Infinium MethylationEPIC v2.0 BeadChips (Illumina, 20087708), in accordance with the manufacturer’s protocol. Patients and normal controls were balanced across 50 arrays and within each array row to reduce technical biases. Patients and normal controls were matched overall for age at sampling to minimize potential confounding effects. DNA methylation arrays were generated at the ASGARD-Rouen genomic platform (University of Rouen and Rouen University Hospital) on an Illumina NextSeq 550 scanner. Raw IDAT data were processed and normalized using the default Meffil R package protocol along with all other samples included in the 50 arrays to better estimate the variability of methylation signals within and across arrays. The samples were functionally normalized together as advocated in the Meffil documentation, with random effect adjustment on array and sentrix row as well as fixed effect adjustment on the first two PCs, before computing β values.

An epigenome-wide association analysis/differential analysis was performed using normalized β values on the subset of normal controls and patients with pathogenic (P) or or likely pathogenic (LP) dominant or recessive biallelic *RNU2-2* variant carriers. For each CpG site, a multiple linear regression model was fitted to assess methylation differences while adjusting for well-known confounders (age at sampling, sex and predicted blood cell composition) in coherence with standard episignature methodology. However, to address possible heterogeneity between variant effects on methylation levels, we added two coefficients accounting for dominant n.35A>G and biallelic recessive variant specific effects on top of the main differential effect between patients and normal controls, n.4G>A serving as “reference variant” accounted for in the main differential term. For each CpG, regression coefficients and corresponding *P*-values were extracted for these three variables of interest. CpGs were considered significant if they passed both thresholds: *P*-value < 1×10^?5^ and absolute effect size > 0.05 on either of these three coefficients. Significant CpGs were further classified according to their association pattern. Indeed, with this modelling strategy, CpGs that share a single common effect across all variant types will have a significant main differential case/control coefficient, but no significant variant-specific coefficient. This set of probes are labelled “probes common to all variants”. In case of heterogeneity, the n.35A>G and biallelic specific coefficients capture the differential effect between the “reference” n.4G>A variant and the variant of interest. The constraint on the minimal average difference of 5% therefore applies to the difference between the average n.4G>A patients and the average variant of interest carriers. Probes reaching significance only at the n.35A>G coefficient are therefore probes where n.4G>A carriers do not significantly or sufficiently (|Δ*β*||>5%) differ on average from normal controls, but n.35A>G carriers are significantly different from n.4G>A carriers. These probes are labelled “probes specific to n.35A>G”. Similarly, probes reaching significance only at the biallelic coefficient were considered “specific to biallelic recessive variant carriers”. In theory, probes that reached significance at the main differential coefficient but also at other variant specific coefficients could correspond to two opposed scenarios: either carriers of the variant of interest display even stronger hyper/hypomethylation than already displayed by n.4G>A carriers or the variant specific effects annul the main differential effect. Visual inspection of the direction of effects at each of these positions confirmed that the second scenario prevailed in all cases. As a consequence, this last set of probes could be considered as “n.4G>A specific probes”.

After filtering, adjusted methylation levels were visualized through PCA and heatmap representations (pheatmap package with the Euclidean distance and Ward aggregation method). In particular, a model adjusting for age at sampling, sex and predicted blood cell composition was fitted on normal control samples for each probe to produce a baseline methylation model. Adjusted methylation levels were computed for each sample from this model by correcting each normalized b value by the expected baseline level among normal controls of similar age, sex and blood composition according to this model. To visualize the distribution of methylation levels across variant groups, CpGs were first classified into categories based on their association patterns in the EWAS (probes common to all variants, n.35A>G specific probes, biallelic recessive specific probes, and n.4G>A specific probes). For each category, the average adjusted methylation level was computed per sample across all CpGs belonging to the corresponding category to summarize the methylation profiles. Raincloud plots were then generated using the ggplot2 and ggdist packages. Each plot combined a half-violin distribution, a boxplot, and individual sample jittered points, with colours assigned to phenotypic groups (controls, dominant n.4G>A, dominant n.35A>G, and recessive biallelic variants). Pairwise group comparisons were performed using two-sided Wilcoxon rank-sum tests implemented in ggpubr, with *P*-values displayed on the plots (non-significant values were hidden).

Finally, to evaluate the predictive performance and the robustness of the CpG signature, we applied a multiclass Support Vector Machine (SVM) classifier with a linear kernel, implemented in the e1071 package in R, with four possible outputs: Control, Dominant n.4G>A, Dominant n.35A>G, Recessive biallelic. To maximize the possible gap between training and testing while keeping reasonable training set sizes, a three-block cross-validation strategy was used: within each variant group, samples were randomly assigned to one of three blocks to maintain class balance across all iterations. At each iteration, two blocks were used for training while the remaining block was kept aside for testing, so that every sample was predicted once. The classifier was trained on the adjusted methylation levels restricted to the selected CpGs, with the outcome defined as subject type (Control, Dominant n.4G>A, Dominant n.35A>G, Recessive biallelic). The SVM model produced predicted class labels and class membership probabilities for each test sample. Performance metrics were calculated per class, including sensitivity and specificity, with 95% confidence intervals estimated using the Wilson method for binomial proportions. Predicted probabilities were averaged across samples and visualized on a scatter plot to illustrate class separation and model confidence.

---

**Extended Data Figure 1.**
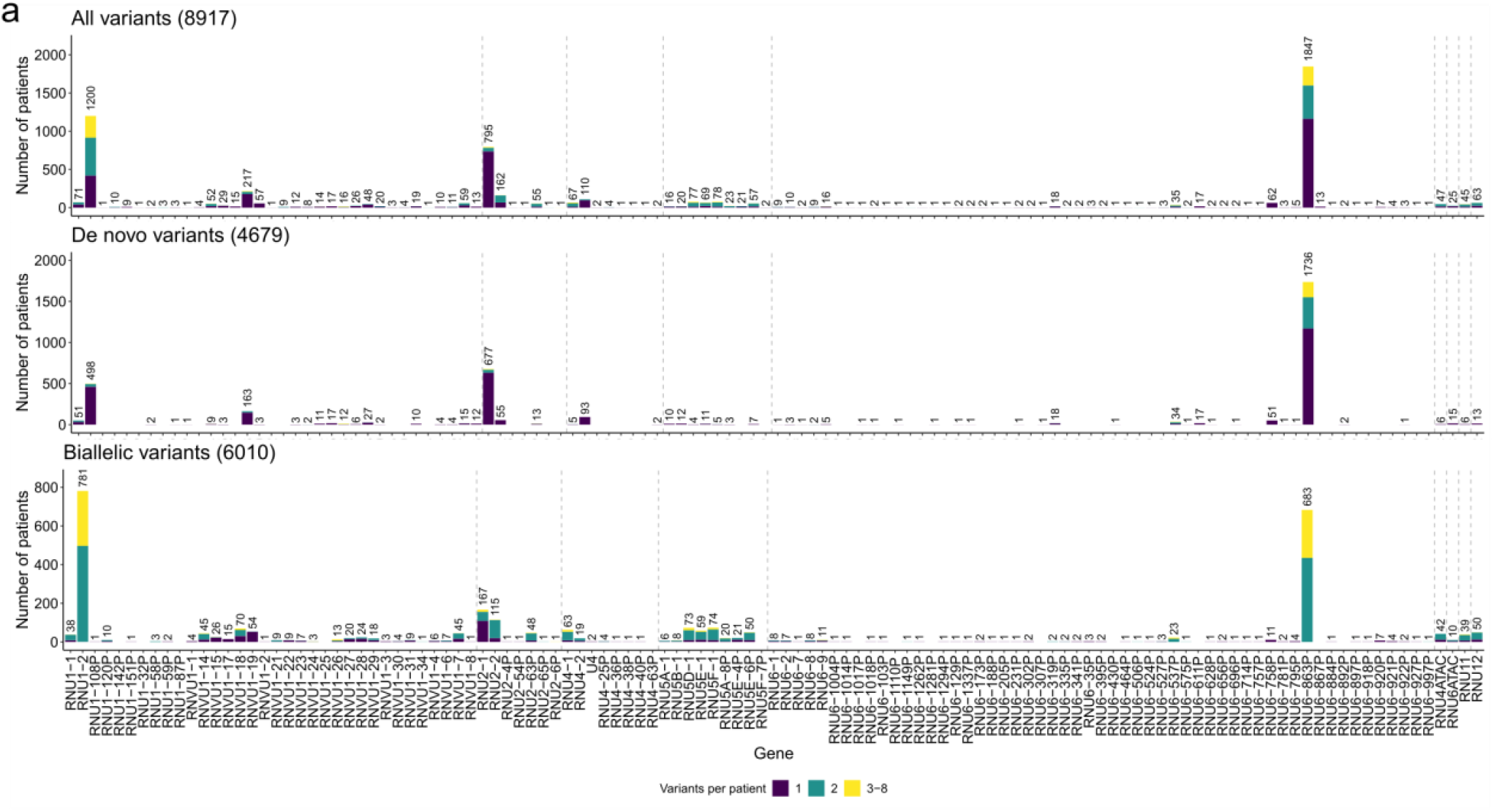
Distribution of variants across 200 putatively functional genes in the PFMG cohort. Supplementary Fig 1 shows the same distribution in the SeqOIA and Auragen subcohorts separately.

**Extended Data Figure 2.**
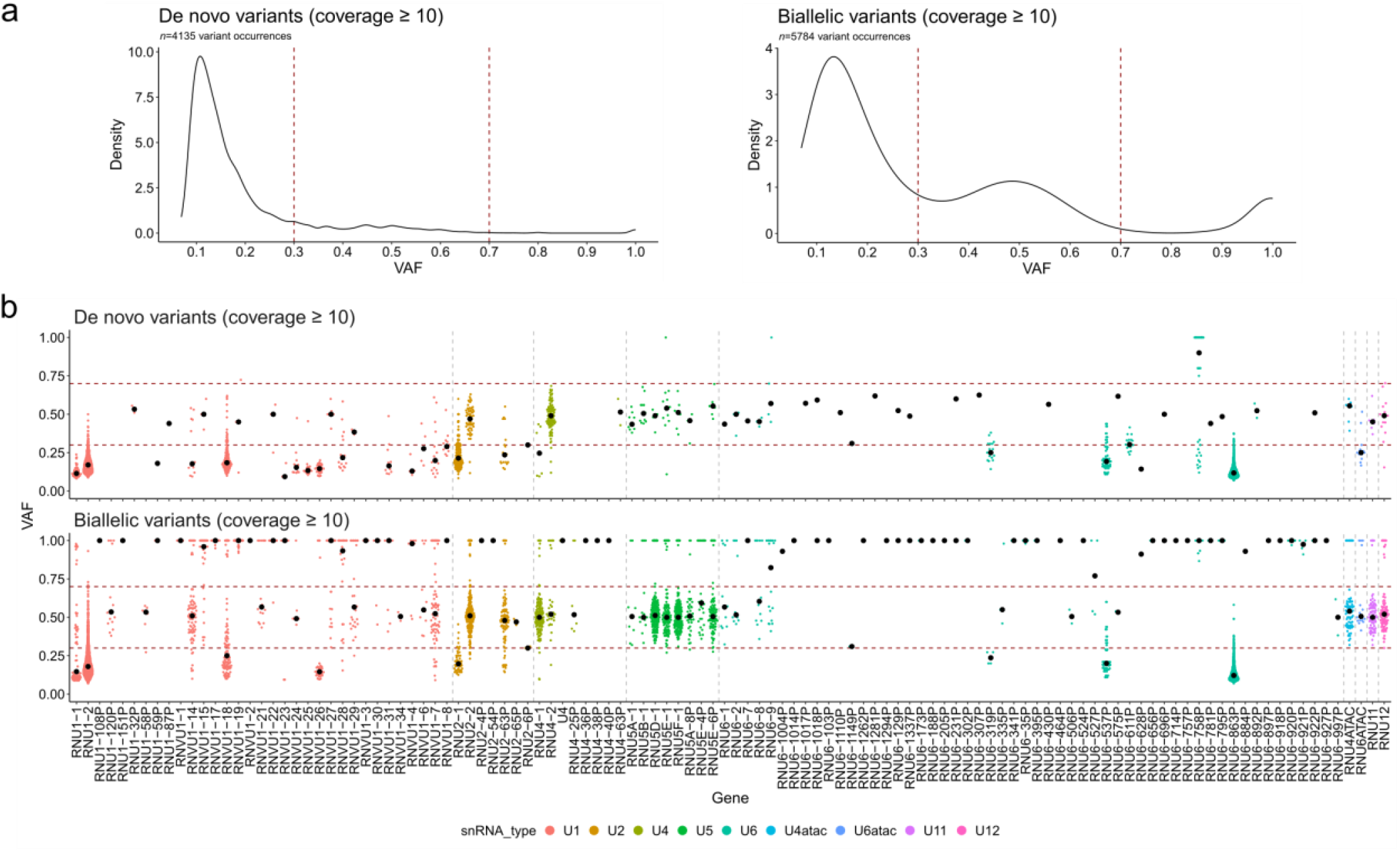
Variant allele fraction (VAF) distributions of snRNA variants in the aggregated PFMG cohort. **a**, VAF distribution of *de novo* variants in snRNA genes with coverage ≥10. **b**, VAF distribution of biallelic variants in snRNA genes with coverage ≥10. **c**, Detailed VAF distribution for snRNA genes harbouring variants in the PFMG cohort. Supplementary Fig 2 shows the same data in the seqOIA and Auragen subcohorts separately.

**Extended Data Figure 3.**
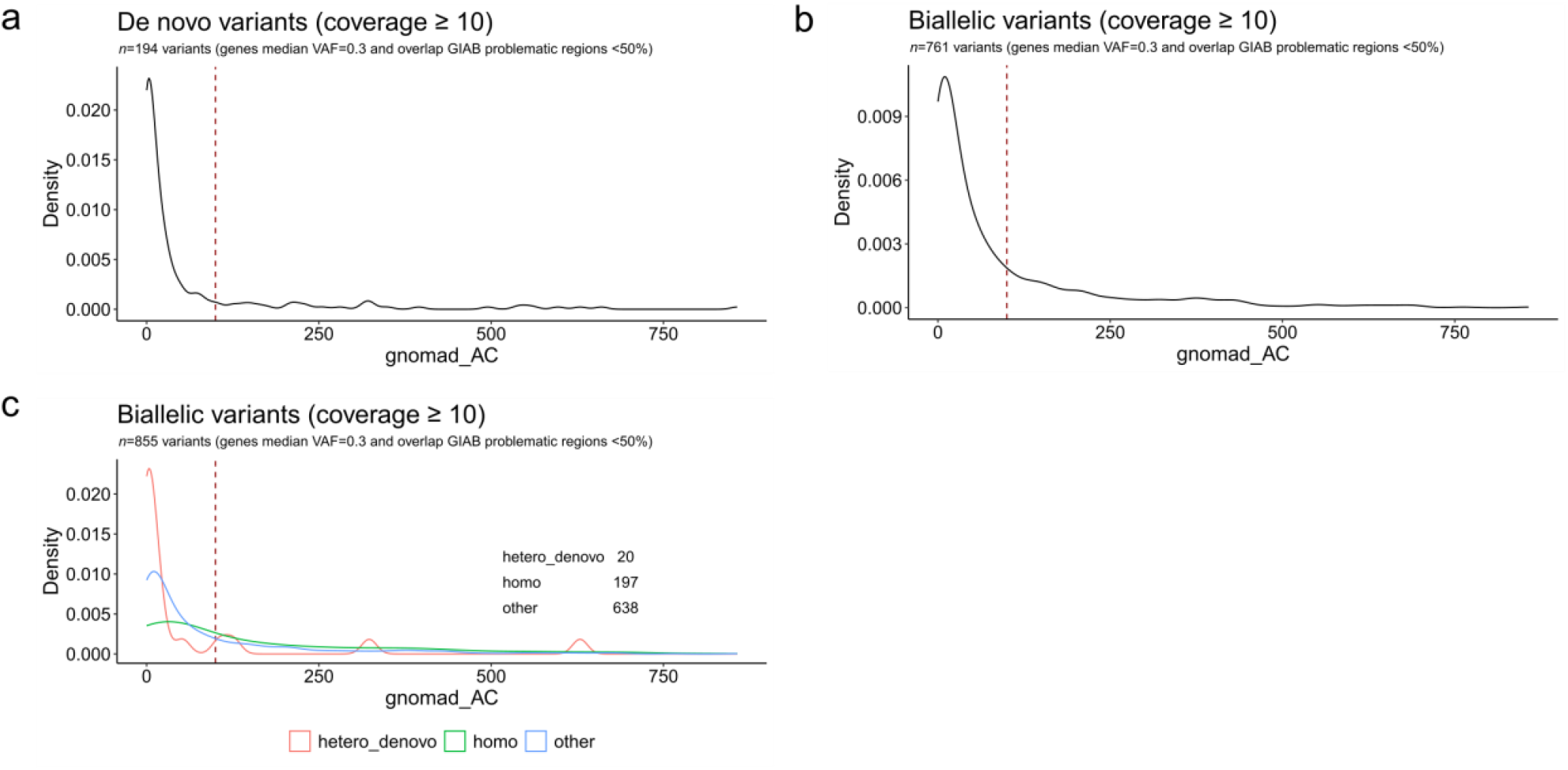
gnomAD allele count (AC) distribution of snRNA variants in the aggregated PFMG cohort. **a**, *De novo* variants with coverage ≥10. **b and c**, Biallelic variants with coverage ≥10 altogether (**b**) or stratified according to their inheritance (**c**). Supplementary Fig 3 shows the same data in the SeqOIA and Auragen subcohorts separately.

**Extended Data Figure 4.**
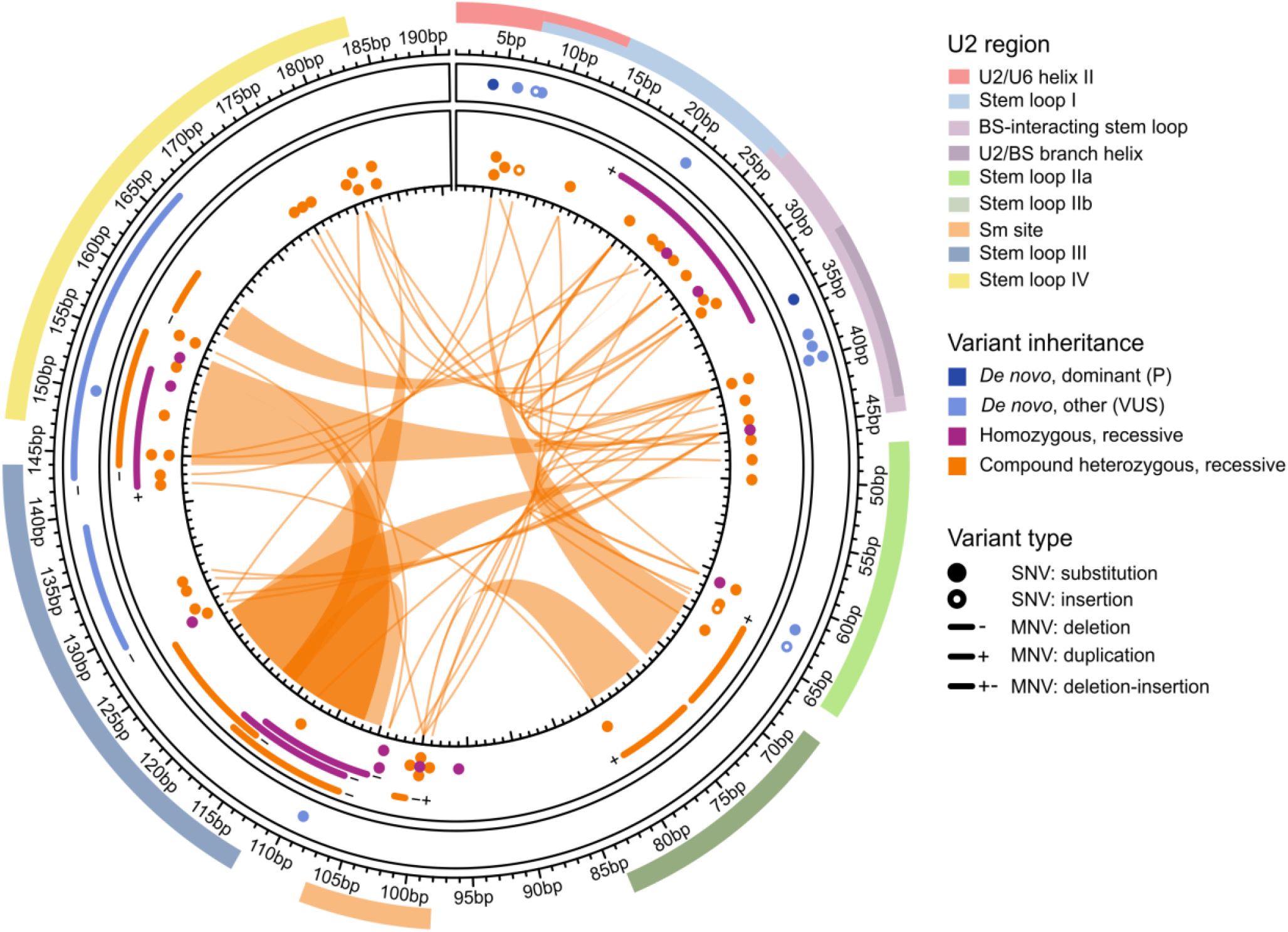
Circos plot depicting preferential associations of biallelic variants. This scheme suggests that, in affected individuals with compound heterozygote variants, variants preferentially co-occur with one affecting the 5′ domain and the other the 3′ domain, suggesting domain-specific combinatorial effects. Variant colouring is the same as in Fig 3a.

**Extended Data Figure 5.**
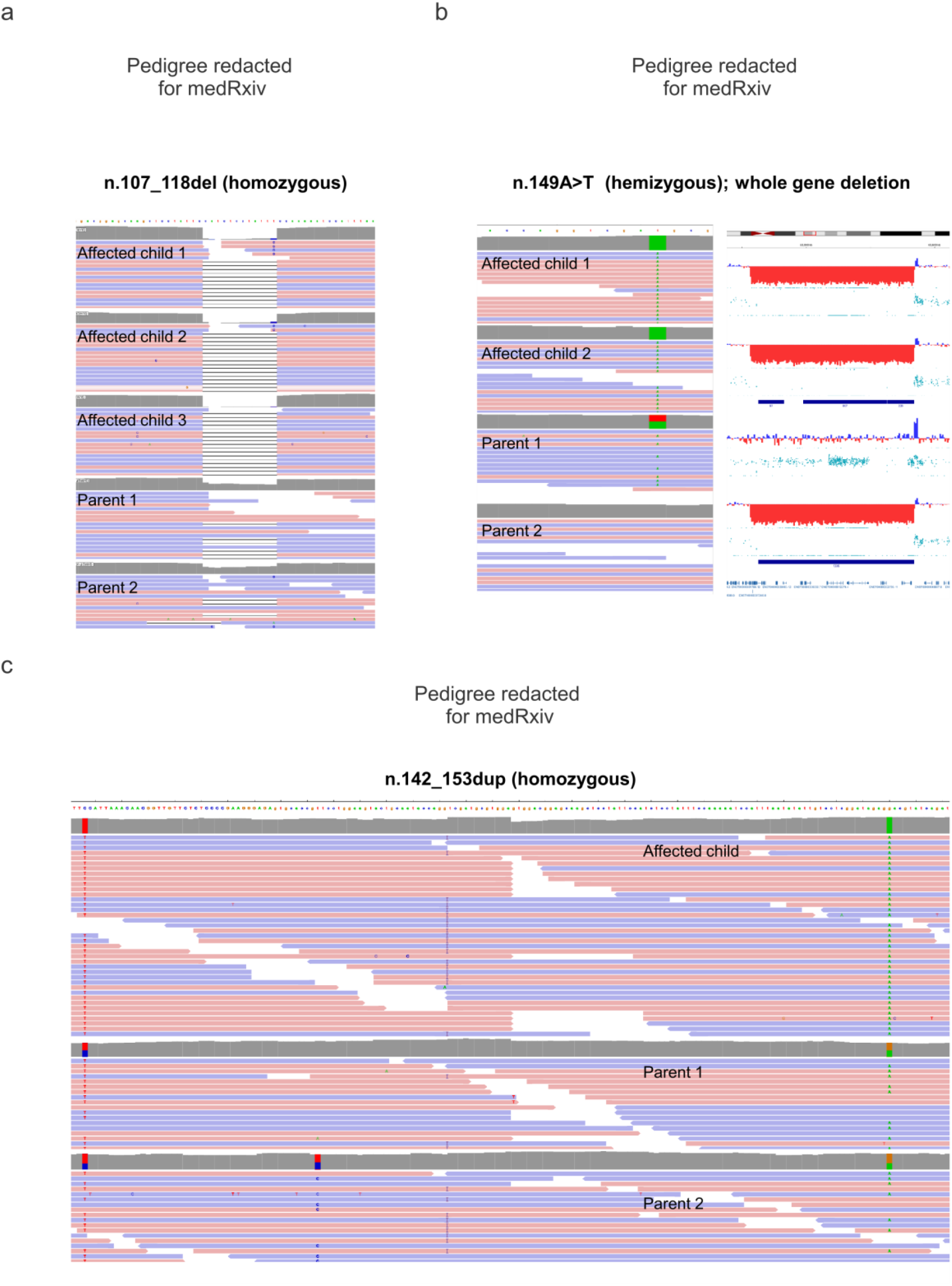
Examples of biallelic variants identified by genome sequencing in the PFMG cohort. **a**, Integrative Genomics Viewer (IGV) screenshots of BAM file alignments showing segregation of the *RNU2-2* n.127_118del variant in the homozygous state in three affected siblings, with both parents carrying the variant in heterozygous state. **b**, Left: IGV screenshot displaying the hemizygous *RNU2-2* n.149A>T variant in two affected siblings and their heterozygous parent. Right: Copy-number analysis of genome sequencing data revealing a 642 kb deletion (hg38: chr11:62,781,800-63,424,493) encompassing *RNU2-2* and 31 additional genes, present in the two affected siblings and the other parent. **c**, IGV screenshot showing the presence of the *RNU2-2* n.142_153dup variant in homozygous state in the affected child and in heterozygous state in both parents.

**Extended Data Figure 6.**
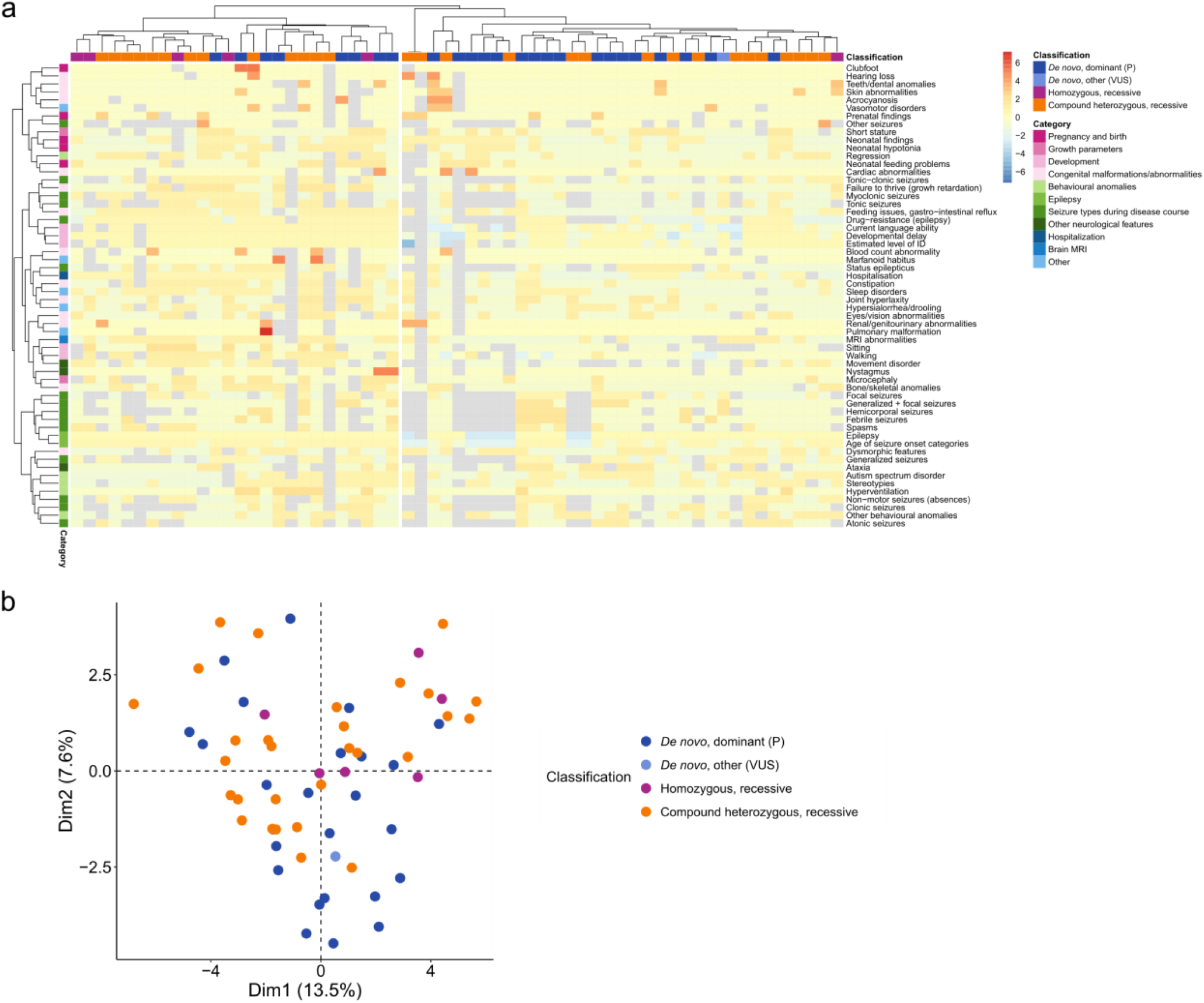
Principal component analysis and classification performance of the *RNU2-2* episignature. **a**, Hierarchical clustering of the clinical features (*n*=58, rows) of patients with *de novo* dominant or biallelic *RNU2-2* variants (*n*=74, columns). Categorical data was converted to 0-1 scale, and values were Z-score scaled for each row. Blue-yellow-red scale depicts Z-scores. Lower values indicate a more favourable phenotype, while higher values represent a more severe phenotype. Missing values are shown in grey. Columns are coloured based on the variant classification: dark blue: *de novo*, dominant (n.4G>A or n.34A>G); light blue: *de novo* other (VUS); orange: compound, recessive; purple: homozygous, recessive. **b**, Principal component analysis of clinical features in *RNU2-2* variant carriers. Missing values were imputed as 0. Variant colouring is the same as in Ext Fig 6a.

**Extended Data Figure 7.**
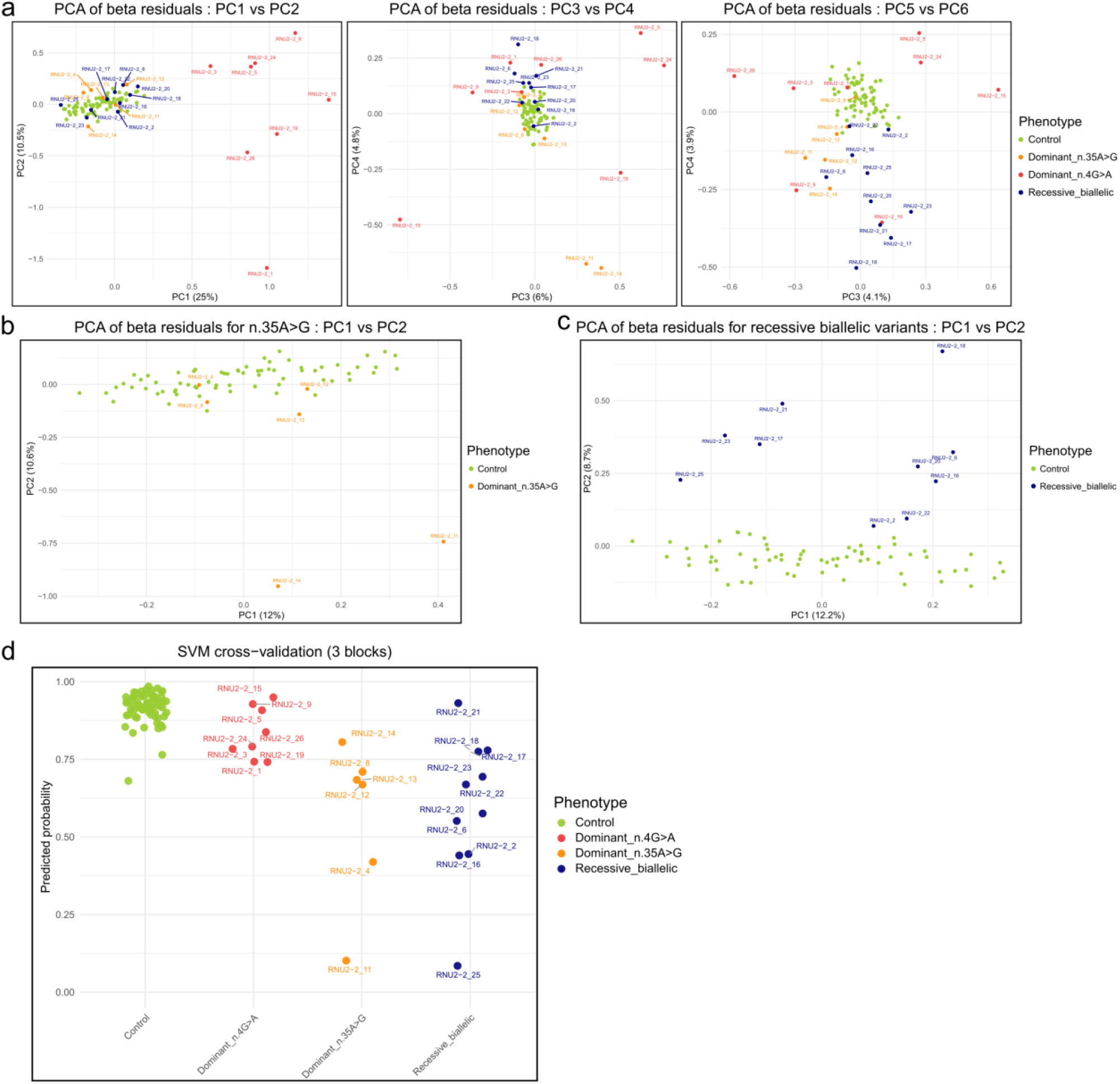
Principal component analysis and classification performance of the *RNU2-2* episignature. In all panels, controls are shown in green, dominant n.4G>A carriers in red, dominant n.35A>G carriers in orange, and recessive biallelic cases in blue. **a**, PCA of adjusted methylation levels at differentially methylated positions (*n*=201) up to component 6, after correction for expected baseline methylation level based on age at sampling, sex and estimated blood cell counts (*n*=92 individuals in total) showing separation of dominant n.4G>A carriers from normal controls on axis 1, and the weaker separation between dominant n.35A>G carriers or recessive biallelic cases from normal controls along axis 6. The percentage of variance explained is provided for each component within the axis title. **b**, PCA of adjusted methylation levels at differentially methylated positions (*n*=201), after correction for expected baseline methylation level based on age at sampling, sex and estimated blood cell counts on the restriction to normal controls and carriers of the dominant n.35A>G variant along the first two principal components (*n*=74 individuals in total). The percentage of variance explained is provided for each axis. **c**, PCA of adjusted methylation levels at differentially methylated positions (*n*=201), after correction for expected baseline methylation level based on age at sampling, sex and estimated blood cell counts on the restriction to normal controls and carriers of recessive biallelic variants along the first two principal components (*n*=78 individuals in total). The percentage of variance explained is provided for each axis. **d**, Predicted probabilities from a three-block cross-validation using a four-class SVM classifier (control, dominant n.4G>A, dominant n.35A>G, recessive biallelic).

**Supplementary Figure 1.**
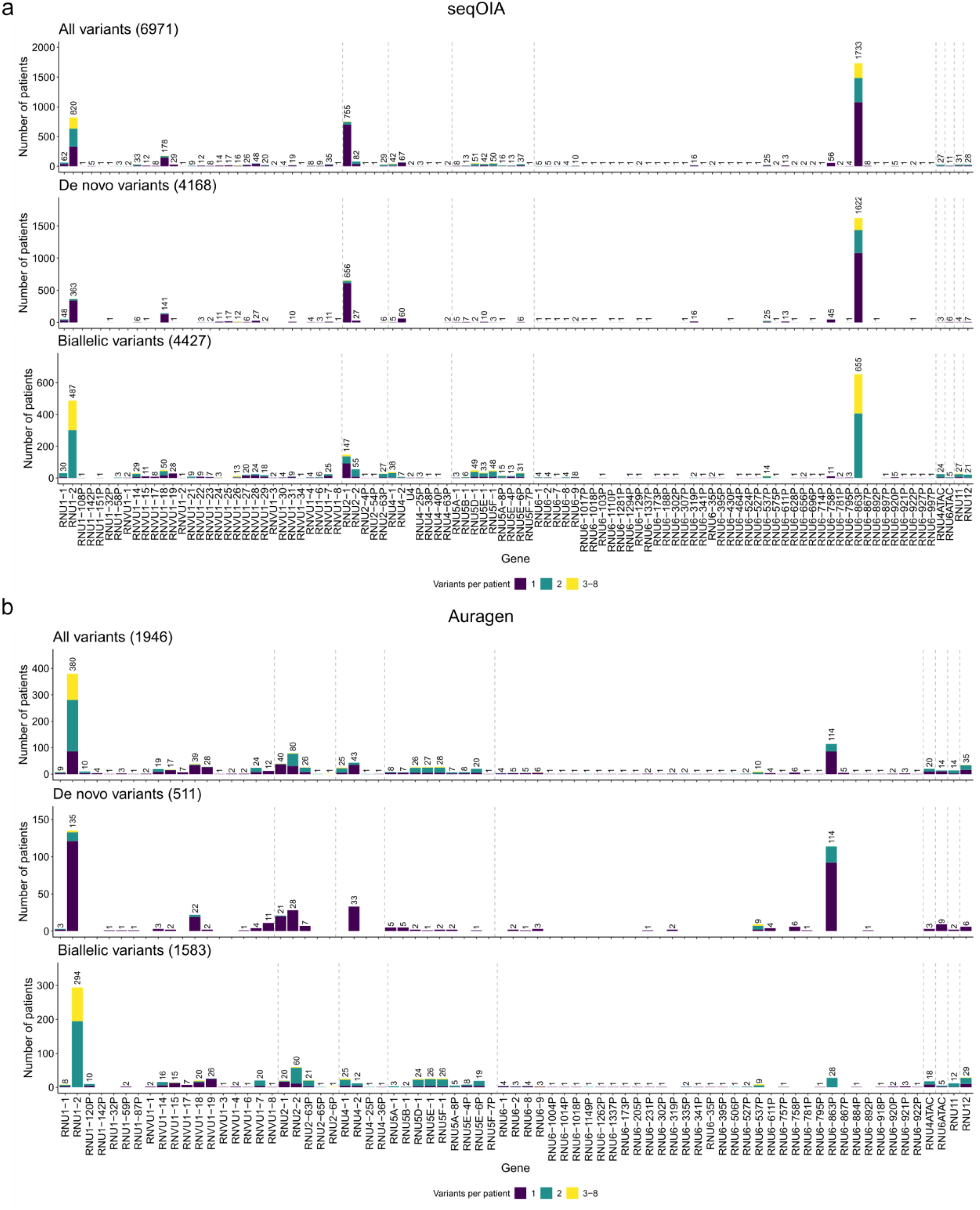
Variant distribution across 200 putatively functional genes in the Auragen and SeqOIA PFMG subcohorts. **a**, SeqIOA; **b**, Auragen.

**Supplementary Figure 2.**
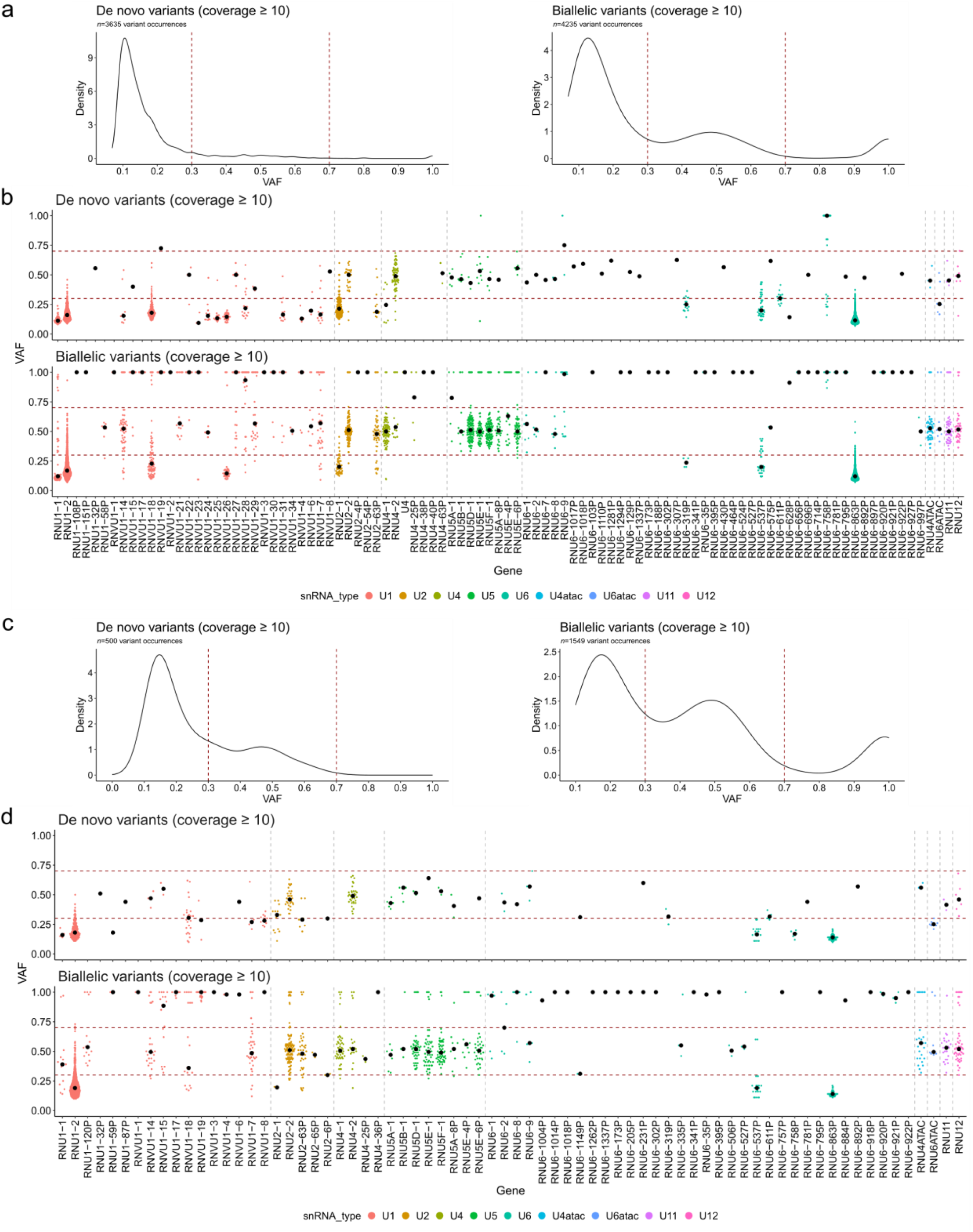
Variant allele fraction (VAF) distributions of snRNA variants in SeqIOA and Auragen subcohorts. **a**, VAF distribution of *de novo* (left panel) or biallelic (right panel) variants in snRNA genes with coverage ≥10 in SeqOIA. **b**, Detailed VAF distribution in SeqOIA. **c**, VAF distribution of *de novo* (left panel) or biallelic (eight panel) variants in snRNA genes with coverage ≥10 in Auragen. **d**, Detailed VAF distribution in Auragen.

**Supplementary Figure 3.**
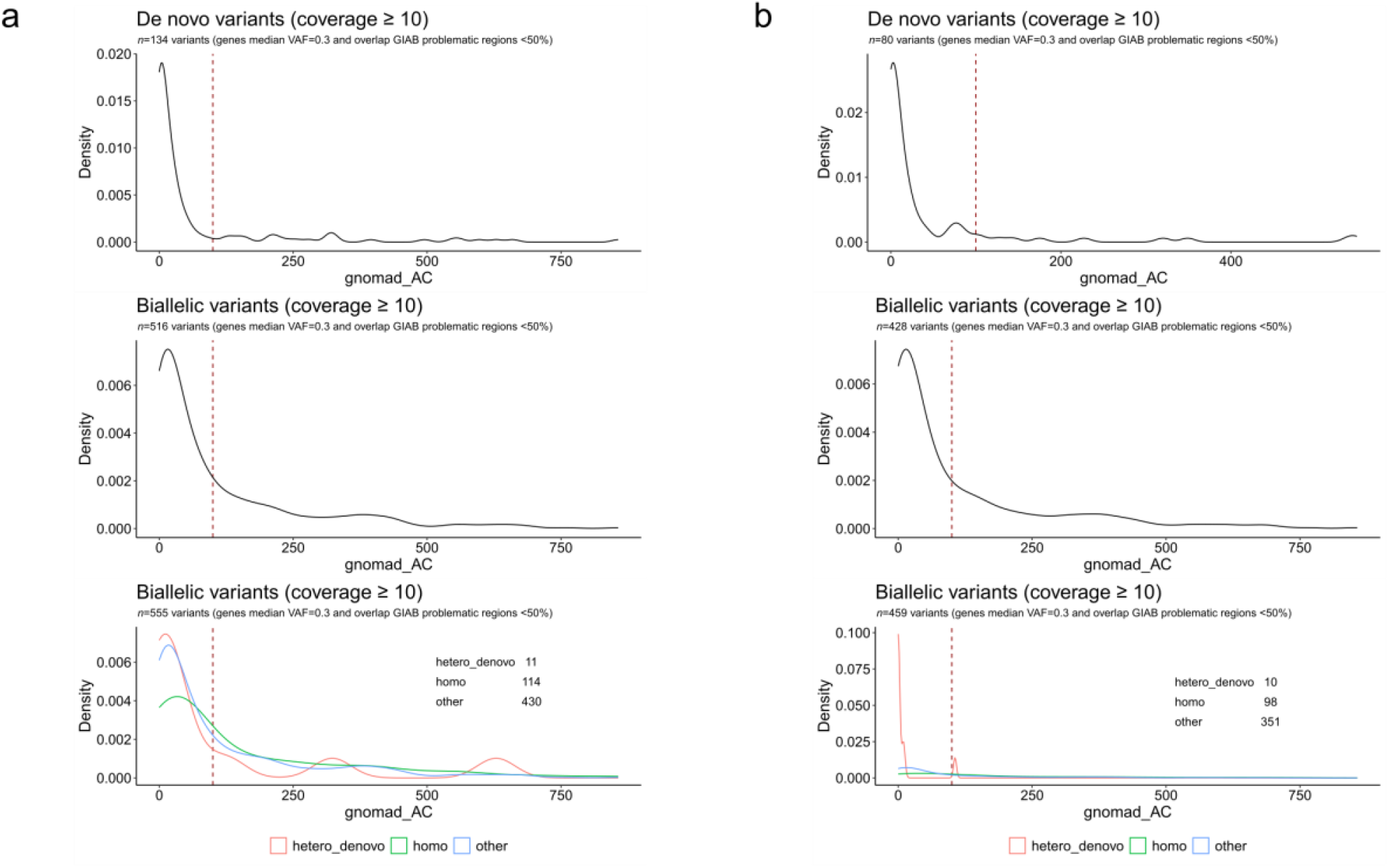
Extended Data Fig 3. gnomAD allele count (AC) distribution of snRNA variants variants in SeqIOA and Auragen subcohorts. **a**, SeqIOA; **b**, Auragen. Upper panel: *de novo* variants with coverage ≥10. Middle and lower panels: biallelic variants with coverage ≥10 altogether (middle) or stratified according to their inheritance (lower).

**Supplementary Figure 4.**
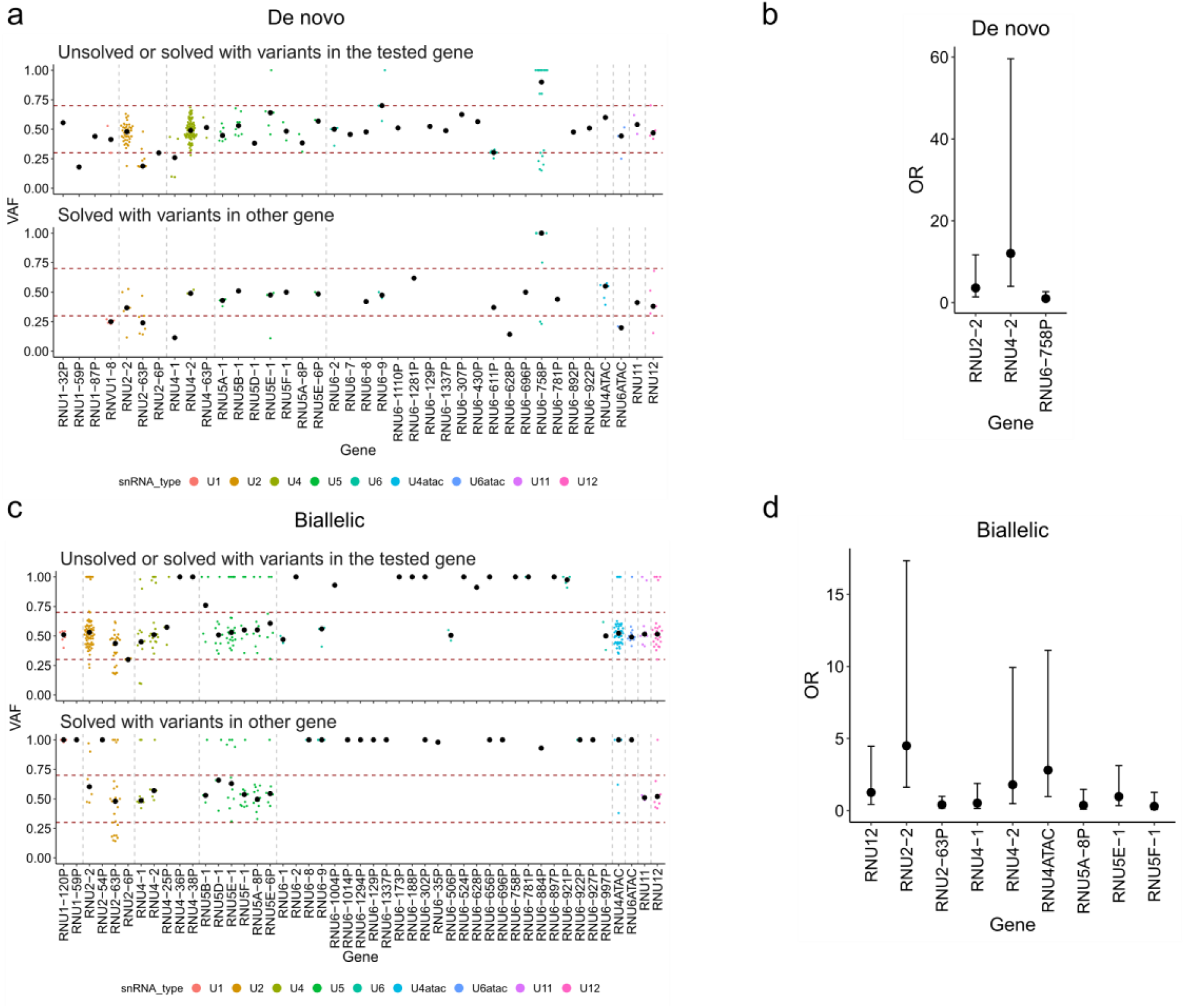
Identification of potential novel snRNA gene-disease associations in the PFMG cohort. The cohort was divided into solved (*n*=8,343) and unsolved (*n*=18,568) cases for discovery analyses, with cases solved by variants in snRNAs with known disease association merged into the unsolved group. We compared the proportion of cases with rare variants (gnomAD allele count < 100) between solved and unsolved groups. **a**-**c**, Variant allele fraction (VAF) distribution for rare *de novo* variants (**a**) and rare biallelic variants (**c**); **b**-**d**, Odds-ratio (OR; dot) and OR confidence interval of unsolved versus solved cases for genes in which at least 10 patients had variants (minimum number needed to reach statistical significance in the cohort); **b**, de novo variants; **d**, biallelic variants. Fisher’s test *P*-values are shown in Fig 2. Supplementary Fig 5 shows the same analyses performed separately in the SeqIOA and Auragen subcohorts.

**Supplementary Figure 5.**
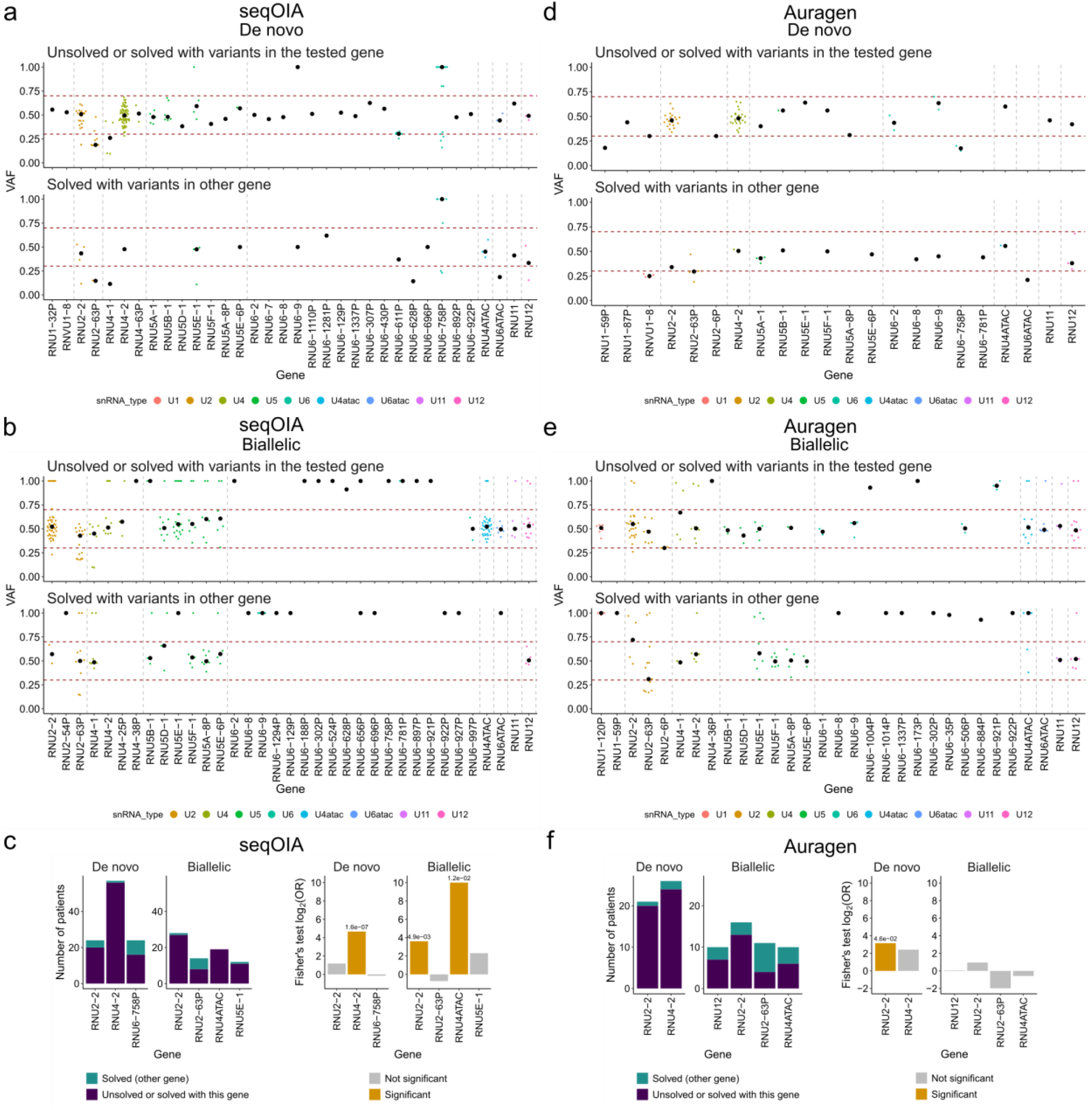
Comparison of rare variant burden in snRNA genes in unsolved versus solved cases separating the PFMG cohort into SeqOIa and Auragen subparts. **a**, and **d**, comparison for rare (gnomAD allele count < 100) *de novo* variants in SeqOIA (a) and Auragen (b); **b**, and **e**, comparison for rare (gnomAD allele count < 100) biallelic variants in SeqOIA (b) and Auragen (e). **c**, and **f**, Number of patients with rare *de novo* and biallelic variants (left panels) and statistical enrichment in unsolved versus unsolved cases (right panels) for genes in which at least 10 patients had variants (minimum number needed to reach statistical significance in the cohort) in SeqOIA (**c**) and Auragen (**f**).

**Supplementary Figure 6.**
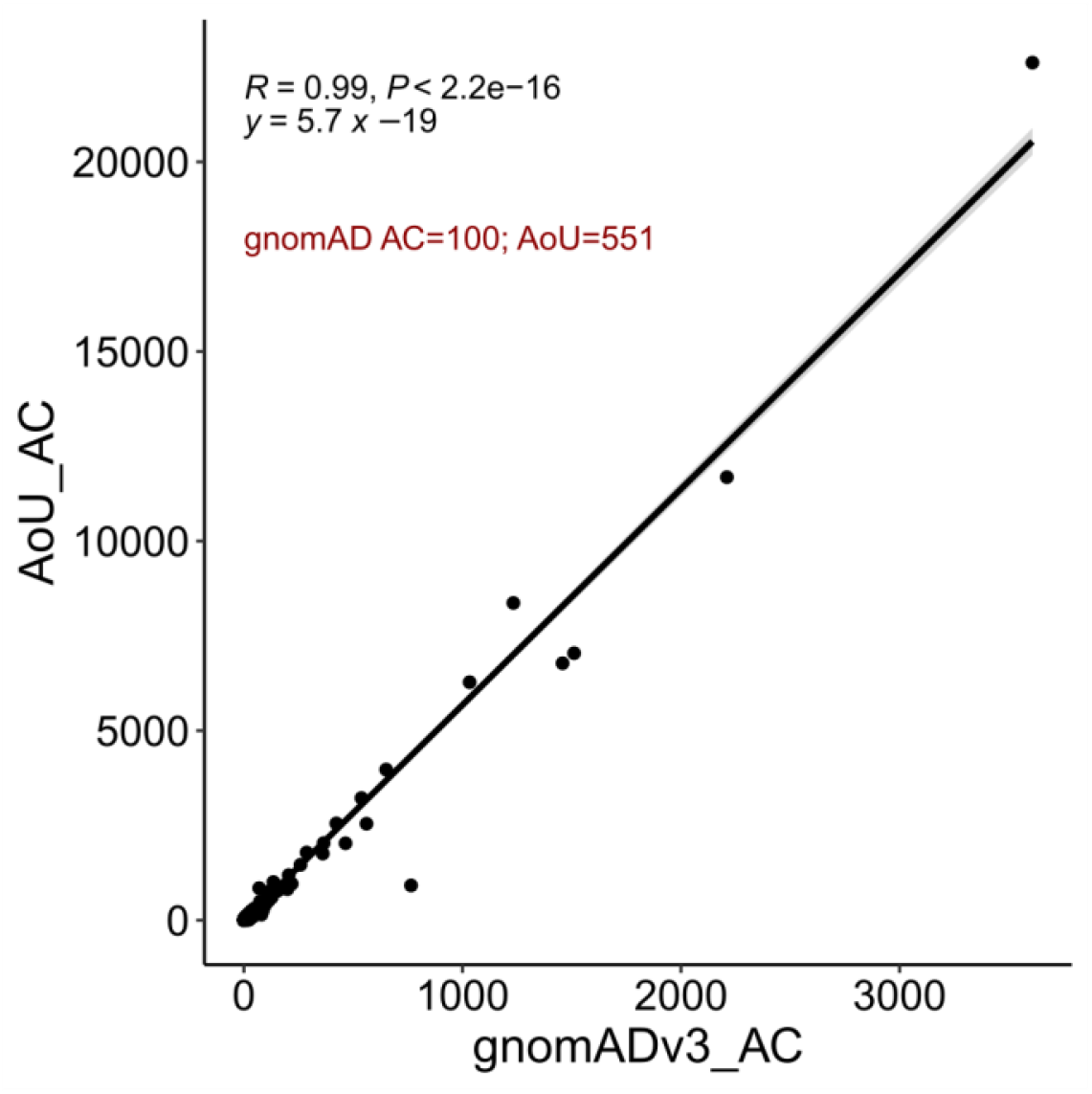
Correlation between gnomAD v3 and *All of Us* allele counts for *RNU2-2* variants. The cut-off of 100 allele counts (AC) in gnomAD v3.1 (76,156 genomes) corresponds to 551 in the *All of Us* database (414,000 genomes). For the PFMG cohort, we applied stricter thresholds: AC<50 for de novo variants and AC<200 for biallelic variants, representing 10× and 2.5× greater stringency, respectively, compared to the gnomAD cut-off.

